# Social determinants of recovery from ongoing symptoms following COVID-19 in two UK longitudinal studies: a prospective cohort study

**DOI:** 10.1101/2023.12.21.23300125

**Authors:** Nathan J. Cheetham, Vicky Bowyer, María Paz García, Ruth C. E. Bowyer, J. D. Carpentieri, Andy Guise, Ellen J. Thompson, Carole H. Sudre, Erika Molteni, Michela Antonelli, Rose S. Penfold, Nicholas R. Harvey, Liane S. Canas, Khaled Rjoob, Benjamin Murray, Eric Kerfoot, The COVID Symptom Study Biobank Consortium, Alexander Hammers, Emma L. Duncan, Claire J. Steves

**Affiliations:** Department of Twin Research and Genetic Epidemiology, King’s College London, London, United Kingdom; The Alan Turing Institute, London, United Kingdom; Institute of Education, University College London, London, United Kingdom; Department of Population Health Sciences, King’s College London, London, United Kingdom; School of Psychology, University of Sussex, Falmer, United Kingdom; MRC Unit for Lifelong Health and Ageing, Department of Population Science and Experimental Medicine, University College London, London, United Kingdom; Centre for Medical Image Computing, Department of Computer Science, University College London, London, United Kingdom; School of Biomedical Engineering & Imaging Sciences, King’s College London, London, UK; Edinburgh Delirium Research Group, Ageing and Health, Usher Institute, University of Edinburgh, Edinburgh, United Kingdom; King’s College London & Guy’s and St Thomas’ PET Centre, King’s College London, London, United Kingdom; Guy’s & St Thomas’s NHS Foundation Trust, London, United Kingdom

**Keywords:** Long COVID, SARS-CoV-2, recovery, ongoing symptoms, social gradient, socio-demographics, health disparities, health inequalities

## Abstract

**Background:** Social gradients in COVID-19 exposure, illness severity, and mortality have been observed in multiple international contexts. Whether pre-existing social factors affect recovery from ongoing symptoms following COVID-19 and long COVID is less well understood.

**Methods:** We analysed data on self-perceived recovery following self-reported COVID-19 illness in two United Kingdom community-based cohorts, COVID Symptom Study Biobank (CSSB) (N = 2548) and TwinsUK (N = 1334). Composite variables quantifying socio-demographic advantage and disadvantage prior to the COVID-19 pandemic were generated from sex, ethnic group, education, local area deprivation and employment status. Associations between self-perceived recovery and composite variables were tested with multivariable logistic regression models weighted for inverse probability of study participation, adjusting for potential confounding by age, region and pre-pandemic health factors, and potential mediation by COVID-19 illness characteristics and adverse experiences during the pandemic. Further analyses tested associations between recovery and individual socio-demographic variables reflecting status prior to and during the COVID-19 pandemic.

**Findings:** Socio-demographic gradients in recovery were observed, with unadjusted recovery rate varying between 50% and 80% in CSSB and 70% and 90% in TwinsUK based on composite socio-demographic variables. Likelihood of recovery was lower for individuals with more indicators of pre-pandemic social disadvantage in both cohorts (CSSB: odds ratio, OR = 0.74, 95% confidence interval, CI: 0.62-0.88, TwinsUK: OR = 0.79, 95% CI: 0.64-0.98 per disadvantage) and higher with more social advantages (CSSB: OR = 1.26, 95% CI: 1.08-1.47, TwinsUK: OR = 1.36, 95% CI: 1.09-1.70 per advantage). Associations were neither explained by differences in COVID-19 illness severity or timing, nor adverse social experiences during the pandemic, which were themselves inversely associated with recovery.

**Interpretation:** Strong social inequalities in the likelihood of recovery from COVID-19 were observed, with ongoing symptoms several months after coronavirus infection more likely for individuals with multiple indicators of social disadvantage. Work is needed to identify modifiable biopsychosocial factors to enable interventions that address inequalities.

**Funding:** Chronic Disease Research Foundation, National Institute for Health and Care Research, Medical Research Council, Wellcome LEAP, Wellcome Trust, Engineering & Physical Sciences Research Council, Biotechnology and Biological Sciences Research Council, Versus Arthritis, European Commission, Zoe Ltd.

**Plain language summary:** Across the world acute COVID-19 illness has affected the most disadvantaged in society the most. However, we have not looked in detail whether people’s social circumstances affect their recovery from COVID-19. In our study, we asked people from two UK-based health studies if they still had symptoms after having COVID-19. We looked at how advantaged or disadvantaged they were at the start of the pandemic, based on information about their sex, ethnic group, education level, local area, and employment. In both studies, people who were more disadvantaged were more likely to still have symptoms long after having COVID-19. In contrast, more advantaged people were more likely to have fully recovered. We also saw that people who had negative experiences during the pandemic such as losing their job, being unable to afford their bills or not being able to access health & social care services were less likely to recover. More work is needed to understand how and why recovery was so different for people with different circumstances.

**Research in context:** *Evidence before this study:* To search for previous reports on associations between recovery from COVID-19 and socio-demographic factors, we screened abstracts identified from the PubMed search query on December 21, 2023: “((COVID-19) AND ((recovery) OR (convalescence) OR (“ ongoing symptoms”)) AND ((socioeconomic) OR (sociodemographic) OR (social) OR (gradient))) AND LitCLONGCOVID[filter]”, where LitCLONGCOVID is a filter for articles relating to long COVID (https://pubmed.ncbi.nlm.nih.gov/help/#covid19-article-filters), which returned 210 results published between July, 2020 and December, 2023. A small number (N = 11) of studies contained direct measures of recovery from COVID-19 in terms of presence/absence of ongoing symptoms relating to COVID-19 illness, either as perceived by the individual or inferred from current symptom reports. Of these, most focused on associations with COVID-19 illness factors such as severity and symptomatology, and prior health indicators. Socio-demographics were mostly used for sample description and adjustments in models rather than as exposures of interest. Of the few studies (N = 8) that tested associations with socio-demographic variables, the range of socio-demographics tested was limited and/or follow-up time typically restricted to 6-12 months since symptom onset. In these studies, associations with recovery were reported for age (N = 4), sex (N = 7), race/ethnicity (N = 2), local area deprivation (N = 1), and education level (N = 1). Associations between long-term symptoms and education or income have been reported in single separate studies. Monthly bulletins up to March 2023 from the UK Coronavirus Infection Survey highlighted prevalence of individuals reporting current effects on daily activities due to long COVID was associated with age, sex, race/ethnicity, local area deprivation and economic activity. No studies were identified that tested for associations of multiple socio-demographics in combination with the likelihood of recovery following COVID-19.

*Added value of this study:* This is the first study to testing the effects of multiple socio-demographics on self-perceived recovery in combination. Measures that attempt to quantify social advantage and disadvantage were generated from multiple known social determinants of health. We tested a wider range of socio-demographic factors than previous studies, including UK geographic region, educational qualification level, employment status and income. Our study has a longer follow-up time than previous comparable reports, with most participants assessed more than one year after infection onset. Detailed data on health before the coronavirus pandemic and COVID-19 illness allowed models to be adjusted extensively and mediation effects to be tested.

*Implications of all the available evidence:* The likelihood of full recovery following COVID-19 appears to follow a social gradient, higher for individuals with multiple indicators of social advantages and fewer disadvantages, and lower for those with multiple social disadvantages and fewer advantages prior to the coronavirus pandemic. This reflects and reaffirms the established cycle of social inequalities in health, between individuals’ status within social hierarchies and ill-health. More work is needed to understand the pathways through which this inequality operates so that interventions can be made.

## Introduction

Following infection with severe acute respiratory syndrome coronavirus 2 (SARS-CoV-2), some individuals report persistent symptoms for months or years [1]. Such individuals may self-identify under the collective patient-advocated term “long COVID” [2], and/or meet one of the various clinical definitions created to describe persistent symptoms [3–5]. The rate of full recovery from ongoing post-COVID-19 symptoms has been reported to be low among individuals with severe acute infection and/or long-term symptoms, with estimates varying between 15% and 50% at up to 12 months from infection [6–10]. Nationally representative estimates from the UK Coronavirus Infection Survey (CIS) estimated 1.9 million individuals (2.8% of UK population) as having self-reported long COVID as of March, 2023 and 1.0 million (1.4%) and 361,000 (0.5%) reported the impact of ongoing symptoms on their current daily activities as “a little” or “a lot”, respectively [11]. To-date, “recovery” following COVID-19 has generally been defined as the absence of ongoing symptoms related to COVID-19, and has been assessed through self-report survey, or inferred from self-report and/or clinical assessment of ongoing symptoms.

While previous studies have investigated associations with recovery rate, most have primarily focused on COVID-19 symptoms and pre-existing health, with few studies examining the effects of socio-demographic characteristics as exposures. The few studies that have looked at this found lower likelihood of recovery for those with lower educational qualification levels [12], living in higher deprivation areas [7] and for female sex [7,8,10,13–15], and conflicting trends with age [7,8,13,14] and race/ethnicity [7,8], typically at up to 12 months follow-up. Further studies in Germany and UK assessing ongoing post-COVID symptoms or illness severity rather than self-perceived recovery directly have also found protective effects against ongoing symptoms for those with higher educational qualification levels [16], higher income [17], and those employed and economically active [11]. A UK qualitative study also identified a slow recovery process and socio-economic challenges to recovery as themes for individuals living with long COVID [18].

Given the known importance of such social determinants of health in other chronic conditions such type 2 diabetes, chronic obstructive pulmonary disease and cardiovascular disease [19–21], and the implications of ongoing COVID-19 symptoms on daily functioning [11], quality of life [8,22], cognitive impairment [23], and increased health risk [1,24], as well as socio-economic consequences such as ability to work [25,26], it is important to test whether relationships exist between recovery from COVID-19 and socio-demographic factors. In this study, our objective was to examine whether self-perceived recovery from COVID-19 was associated with: (1) measures of multiple social advantage and disadvantage derived on the basis of individuals’ positions within systems of social power and oppression [27]; and (2) individual socio-demographic factors (illustrated in our directed acyclic graph in Figure 1). Our study is motivated by previous work on the intersectionality framework [28,29]. We hypothesised that recovery from COVID-19 is associated with exposure to multiple socio-demographic advantages and disadvantages, with individuals exposed to more forms of advantage more likely to recover and those exposed to more forms of disadvantage less likely to recover. We also hypothesised that any observed relationship between socio-demographic advantage and recovery would be mediated by differential susceptibility to more severe COVID-19 illness.

**Figure 1.**
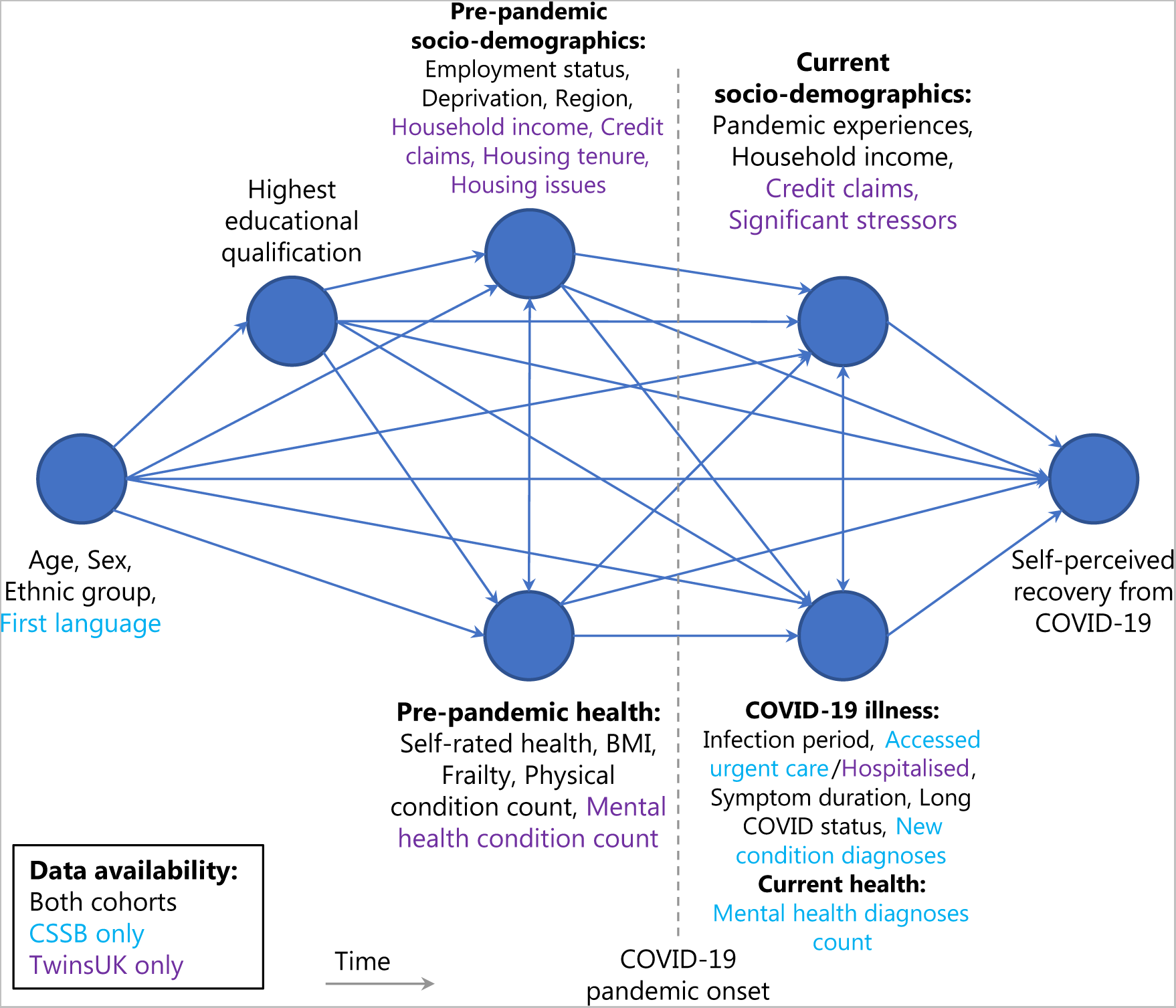
Directed acyclic graph describing hypothesised causal pathways. Proposed directed acyclic graph (DAG) used to generate minimal adjustment variable sets for models estimating the total causal effect of exposure variables on the outcome of self-perceived COVID-19 recovery. The DAG is structured approximately in order of data generation from left to right.

## Methods

### Data sources

Study participants were volunteers from the COVID Symptom Study Biobank (CSSB) and TwinsUK cohorts. Data collection timelines for both cohorts are visualised in Figure S1.

#### COVID Symptom Study Biobank cohort

CSSB participants were recruited via the COVID Symptom Study app from ZOE Ltd (CSS, later renamed ZOE Health Study) launched in the UK on March 24, 2020. All data were collected with informed consent obtained online. Via the CSS app, participants self-report demographic information, symptoms potentially suggestive of COVID-19 infection, any SARS-CoV-2 testing and results, and any vaccinations. CSS participants from across the UK were invited to join the CSSB by email in October to November 2020 and May to June 2021.

CSSB invitation targeted five groups with different statuses at the time of invitation as follows: asymptomatic COVID (positive SARS-CoV-2 test and no associated symptoms); “short COVID” (positive SARS-CoV-2 test and 1-13 days of symptoms); “long COVID” (positive SARS-CoV-2 test and ≥ 28 days’ symptoms); “long non-COVID” (negative SARS-CoV-2 test and ≥ 28 days’ symptoms); and “healthy non-COVID” (negative SARS-CoV-2 test and ≤ 3 days with ≤ 3 symptoms). Before invitation, individuals were matched based on minimum Euclidian distance for age, sex and body mass index (BMI) across groups. Due to this targeted approach designed to give five equally-sized, matched groups, cohort composition is not representative of population prevalence of COVID-19 and long COVID. Further background details of cohort are reported elsewhere [23,30].

CSSB participants were invited (N = 8324) to participate in the “COVID Reflections-Two Years On” online questionnaire in August, 2022. Questionnaire data was supplemented with data collected at time of registration with the CSS app, at consent to CSSB, from an earlier CSS app-based questionnaire on mental health in February, 2021, and in an earlier CSSB online questionnaire (“Effects of the Coronavirus Disease (COVID-19) pandemic on life in the UK”) in May, 2021. Variables described below were collected as part of the August, 2022 questionnaire unless otherwise stated. The CSSB Volunteer Advisory Panel were consulted on the delivery of the August, 2022 questionnaire and gave recommendations on invitation and reminder strategies.

#### TwinsUK cohort

TwinsUK is a UK-based national registry of monozygotic and dizygotic twins, with over 15,000 twins registered since 1992 [31]. During the COVID-19 pandemic, TwinsUK participants were invited (N = 8869) to a series of “COVID-19 personal experience” (CoPE) questionnaires [32]. Responses for each round of the CoPE series were collected as follows: (#1) April-May, 2020, (#2) July-August, 2020, (#3) October-December, 2020, (#4) April-July, 2021, (#5) November, 2021 - February, 2022, and (#6) April-May, 2022. CSSB questionnaires were developed in part from CoPE questionnaires, leading to a high degree of overlap in the types of data collected. COVID-19 questionnaire data were supplemented with data collected as part of TwinsUK routine longitudinal questionnaires both before and during the COVID-19 pandemic.

#### Outcome: Self-perceived recovery

Self-perceived recovery following COVID-19 was measured with a single question in CSSB questionnaires and TwinsUK CoPE rounds #3, #4, #5 and #6: “Thinking about the <u>last</u> or only episode of COVID-19 you have had, have you now recovered and are back to normal?”, with the following response options: “Yes, I am back to normal”, “No, I still have some or all my symptoms”. All participants with self-reported COVID-19 were asked about their COVID-19 recovery, except in questionnaires that asked whether an infection was asymptomatic (CSSB August, 2022 and CoPE #3, #5), where asymptomatic individuals were not asked about recovery. CSSB analyses considered recovery status at the August, 2022 questionnaire only, while TwinsUK analyses took the latest available recovery status from CoPE #3 to #6 for each individual to maximise sample size. Questions relating to long COVID including self-perceived recovery were refined from feedback received by the TwinsUK Volunteer Advisory Panel and the Public Involvement Advisory Group of the “CONVALESCENCE” study of long COVID [33].

#### Socio-demographic, health, and COVID-19 illness characteristics

For CSSB participants, socio-demographic characteristics were measured or derived from self-report at registration to the CSS app, CSSB consent, or in the August, 2022 CSSB questionnaire. Health characteristics were measured or derived from self-report at registration to the CSS app, CSSB consent, in a February, 2021 CSS questionnaire, or in the May, 2021 or August, 2022 CSSB questionnaires. COVID-19 illness characteristics were measured or derived from self-report in the August, 2022 and May, 2021 CSSB questionnaires.

For TwinsUK participants, socio-demographic and health characteristics were measured or derived from self-report in COVID-19 or routine longitudinal questionnaires. COVID-19 illness characteristics were measured or derived from self-report in COVID-19 questionnaires.

Full details of data sources, question wording and processing prior to analysis are given in Supplementary Information Section 1 & 2.

#### Composite socio-demographic advantage and disadvantage variables

To investigate the effects of multiple pre-pandemic socio-demographic factors in combination upon recovery from COVID-19, composite variables were generated which counted the number of positions associated with social advantage and/or disadvantage. Individual variables used to generate the composites were selected after consideration of known systems of social power and existing structural social and health inequalities [34–38], as well as availability of data across both cohorts. Individual variables selected were sex, ethnic group, highest educational qualification, local area deprivation, and pre-pandemic employment status. For each individual variable, certain categories were assigned as indicators of socio-demographic ‘advantage’ or ‘disadvantage’ (Table 1).

**Table 1.** Choice of socio-demographic categories as indicators of socio-demographic advantage and disadvantage.

| **Socio-demographic variable** | **Category indicative of socio-demographic *advantage*** | **Category indicative of socio-demographic *disadvantage*** |
| --- | --- | --- |
| Sex | Male | Female |
| Ethnic group | White groups | Racially minoritised groups (Asian/Asian British groups, Black/Black British groups, Mixed/Multiple ethnic groups, Other ethnic groups) |
| Highest educational qualification | Postgraduate degree or higher | Less than University degree or equivalent |
| Local area deprivation | Index of Multiple Deprivation (IMD) Decile 8-10 (least deprived 30%) | IMD Decile 1-3 (most deprived 30%) |
| Pre-pandemic employment status | N/A (no category assigned as advantage) | Unemployed or not in work for medical reasons |

Three measures were generated, grouping individuals according to the number of indicators of socio-demographic advantage (1), disadvantage (2), or the number of advantages minus the number of disadvantages (3). In sensitivity analyses testing the persistence of associations, sex was removed when generating the composite measures.

No weighting was applied when generating composite variables, effectively treating each advantage or disadvantage as equal. We chose not to weight on the basis of the results of the closest equivalent analysis to date [7,8], where logistic regression coefficients for models testing association between recovery were within approximately 50% of each other in magnitude for sex, ethnicity, and deprivation, while pre-pandemic employment status has not previously been tested.

### Eligibility criteria

CSSB and TwinsUK analyses included all individuals with self-reported COVID-19. For all analyses, inclusion criteria were complete data on age, sex, ethnic group, area of residence, and completion of the COVID-19 recovery question (in the CSSB August, 2022, questionnaire or one or more of the TwinsUK CoPE #3 to #6 questionnaires) as the outcome of interest. Individuals who reported an asymptomatic SARS-CoV-2 infection, or whose longest symptom duration COVID-19 episode started less than 84 days before questionnaire completion, were excluded. A full sample selection flow diagram detailing exclusions is given in Figure S2.

### Statistical analysis

#### Regression models & proposed causal pathways

We used multivariable logistic regression models to obtain estimates of effects of socio-demographic variables on the outcome of recovery following COVID-19, i.e., reporting “Yes, I am back to normal”.

Separate models were run for each exposure variable, including potential confounding variables as appropriate based on the hypothesised directed acyclic graph (DAG) (Figure 1), developed using DAGitty software: http://www.dagitty.net/dags.html (full DAGitty code is available openly on GitHub at https://github.com/nathan-cheetham/CSSBiobank_COVIDRecovery) [39]. Models used the “HC3” estimator of coefficient standard errors to account for heteroskedasticity [40].

Primary analyses tested the effects of multiple pre-pandemic socio-demographic disadvantages and advantages on the COVID-19 recovery outcome. Mediation analyses tested the role of COVID-19 illness characteristics and pandemic experiences in mediating the effects of socio-demographic on COVID-19 recovery, with COVID-19 illness characteristics and pandemic experiences included in models as potential mediators in addition to potential confounders. Secondary analyses tested associations between the COVID-19 recovery outcome and individual pre-pandemic and current socio-demographics characteristics, pre-pandemic health characteristics, and COVID-19 illness characteristics. Models included potential confounding socio-demographic and health variables based on the hypothesised DAG. Sensitivity analysis tested associations between COVID-19 recovery and multiple socio-demographic advantages and disadvantages in CSSB samples stratified by sex and not considering sex in the composite measures.

#### Generation of inverse participation weights

To reduce potential bias from differential response rates, models testing associations with COVID-19 recovery included inverse probability of questionnaire response weights, following methods used in previous CSSB studies [23]. Weights were generated from a multivariable logistic regression model predicting questionnaire response (AUC-ROC scores of 0.82 for CSSB and 0.88 for TwinsUK models). The CSSB model comprised the following variables: age group, sex, ethnic group, CSSB invitation round and recruitment group, number of physical health conditions (at registration with CSS app), PRISMA-7 scale score (at registration with CSS app) [41], region, local area deprivation, BMI, number of mental health conditions (from February, 2021 CSS questionnaire), average PHQ-4 scale mental health assessment score in prior CSSB studies [42], SARS-CoV-2 infection status and associated symptom duration from CSS app prospective logging (considering tests and symptoms up to April 28, 2022 [free SARS-CoV-2 tests were no longer freely available in the UK after April 1, 2022]), and number of non-responses to prior CSSB studies (see [23] for additional details). The TwinsUK model predicted probability of participation in one or more of the CoPE #3 to #6 questionnaires, and comprised the following variables: age group, sex, ethnic group, region, local area deprivation, highest educational qualification, number of non-responses to prior questionnaires, and latest available pre-pandemic data for: household income, BMI, PRISMA-7 score, number of physical health conditions, number of mental health conditions.

#### Software

Analyses were performed using python v3.8.8 and packages: numpy v1.20.1, pandas v1.2.4, statsmodels v0.12.2, scipy v1.6.2, scikit-learn v0.24.1, matplotlib v3.3.4, seaborn v0.11.1.

## Results

### Sample characteristics

Data from 2,548 CSSB participants (of 3,731 questionnaire respondents) and 1,334 TwinsUK participants (of 5,466 questionnaire respondents) with one or more self-reported COVID-19 illnesses were analysed (sample selection shown in Figure S2).

Across both cohorts, median age group was 50-60 years old, most individuals were female sex, identified as white ethnic groups, lived in less deprived areas, and were employed immediately before the COVID-19 pandemic (Table 2). Educational qualification levels among CSSB participants were high in comparison to TwinsUK and the general UK population. General health was self-rated prior to the COVID-19 pandemic as “excellent” or “very good” by just over half in both cohorts (CSSB: 56%, TwinsUK: 58%, from extended sample characteristics, Table S1 and Table S2).

**Table 2.** Sample characteristics. Group size and COVID-19 recovery rate among those with self-reported COVID-19 in CSSB and TwinsUK cohorts.

| Variable | Category | CSSB |  | TwinsUK |  |
| --- | --- | --- | --- | --- | --- |
|  |  | Group size, N (%) | COVID-19 recovery (%) | Group size, N (%) | COVID-19 recovery (%) |
|  | **TOTAL** | **2548** |  | **1334** |  |
| COVID-19 recovery status | Recovered | 1748 (68.6%) | 68.6% | 1078 (80.8%) | 80.8% |
|  | Not recovered | 800 (31.4%) |  | 256 (19.2%) |  |
| Age (years) | Median (IQR) | 58 (51-65) |  | 56 (44-66) |  |
| Age group (years) | 18-39 | 155 (6.1%) | 71.6% | 266 (19.9%) | 88.3% |
|  | 40-49 | 386 (15.1%) | 63.5% | 195 (14.6%) | 79.0% |
|  | 50-59 (reference) | 850 (33.4%) | 65.4% | 338 (25.3%) | 78.1% |
|  | 60-69 | 873 (34.3%) | 71.1% | 294 (22.0%) | 79.3% |
|  | ≥ 70 | 284 (11.1%) | 75.7% | 241 (18.1%) | 79.7% |
| Sex | Female (reference) | 2077 (81.5%) | 67.6% | 1153 (86.4%) | 79.7% |
|  | Male | 471 (18.5%) | 72.8% | 181 (13.6%) | 87.8% |
| Ethnic group | Racially minoritised groups | 72 (2.8%) | 58.3% | 52 (3.9%) | 73.1% |
|  | White groups (reference) | 2476 (97.2%) | 68.9% | 1282 (96.1%) | 81.1% |
| Highest educational qualification | Prefer not to answer/Not stated/Unknown | 102 (4.0%) | 56.9% | 53 (4.0%) | 66.0% |
|  | Less than University degree or equivalent | 701 (27.5%) | 64.8% | 680 (51.0%) | 78.4% |
|  | University degree (reference) | 874 (34.3%) | 70.8% | 386 (28.9%) | 84.5% |
|  | Postgraduate degree or higher | 871 (34.2%) | 70.8% | 215 (16.1%) | 85.6% |
| Pre-pandemic employment status | Employed (reference) | 1360 (53.4%) | 66.0% | 689 (51.6%) | 82.3% |
|  | Self-employed | 294 (11.5%) | 68.7% | 119 (8.9%) | 80.7% |
|  | Unemployed | 11 (0.4%) | 63.6% | 9 (0.7%) | 100.0% |
|  | Permanently or long-term sick or disabled | 32 (1.3%) | 53.1% | 18 (1.3%) | 44.4% |
|  | Retired | 568 (22.3%) | 76.4% | 279 (20.9%) | 80.3% |
|  | Other | 216 (8.5%) | 68.5% | 124 (9.3%) | 77.4% |
|  | Unknown | 67 (2.6%) | 62.7% | 96 (7.2%) | 81.2% |
| Local area deprivation | IMD Decile 1-3 (most deprived 30%) | 299 (11.7%) | 62.2% | 178 (13.3%) | 78.7% |
|  | IMD Decile 4-7 (reference) | 999 (39.2%) | 67.1% | 533 (40.0%) | 81.1% |
|  | IMD Decile 8-10 (least deprived 30%) | 1250 (49.1%) | 71.4% | 623 (46.7%) | 81.2% |
| Region | East Midlands | 151 (5.9%) | 60.3% | 69 (5.2%) | 72.5% |
|  | East of England | 275 (10.8%) | 68.4% | 174 (13.0%) | 83.9% |
|  | London (reference) | 475 (18.6%) | 71.2% | 266 (19.9%) | 83.1% |
|  | North East | 78 (3.1%) | 69.2% | 32 (2.4%) | 78.1% |
|  | North West | 274 (10.8%) | 65.0% | 93 (7.0%) | 86.0% |
|  | Scotland & Northern Ireland | 119 (4.7%) | 63.0% | 45 (3.4%) | 73.3% |
|  | South East | 483 (19.0%) | 72.0% | 310 (23.2%) | 80.0% |
|  | South West | 253 (9.9%) | 71.1% | 142 (10.6%) | 81.0% |
|  | Wales | 115 (4.5%) | 63.5% | 47 (3.5%) | 70.2% |
|  | West Midlands | 158 (6.2%) | 71.5% | 79 (5.9%) | 82.3% |
|  | Yorkshire and The Humber | 167 (6.6%) | 65.9% | 77 (5.8%) | 80.5% |
| Number of disadvantage indicators (of 5) | 0 | 299 (11.7%) | 76.3% | 81 (6.1%) | 90.1% |
|  | 1 (reference) | 1436 (56.4%) | 69.6% | 514 (38.5%) | 82.7% |
|  | 2 | 689 (27.0%) | 65.9% | 620 (46.5%) | 79.7% |
|  | ≥ 3 | 124 (4.9%) | 54.0% | 119 (8.9%) | 72.3% |
| Number of advantage indicators (of 4) | 0 | 15 (0.6%) | 53.3% | 39 (2.9%) | 82.1% |
|  | 1 | 739 (29.0%) | 63.6% | 518 (38.8%) | 78.8% |
|  | 2 (reference) | 1145 (44.9%) | 69.4% | 592 (44.4%) | 79.9% |
|  | 3 (CSSB) / ≥ 3 (TwinsUK) | 557 (21.9%) | 72.2% | 185 (13.9%) | 89.2% |
|  | 4 (CSSB only) | 92 (3.6%) | 79.3% |  |  |
| Number of advantages – number of disadvantages | ≤ -3 | 12 (0.5%) | 58.3% | 30 (2.2%) | 76.7% |
|  | -2 | 115 (4.5%) | 52.2% | 98 (7.3%) | 73.5% |
|  | -1 | 316 (12.4%) | 62.7% | 298 (22.3%) | 78.9% |
|  | 0 | 681 (26.7%) | 69.2% | 448 (33.6%) | 80.4% |
|  | +1 (reference) | 717 (28.1%) | 68.8% | 257 (19.3%) | 80.5% |
|  | 2 | 471 (18.5%) | 71.3% | 141 (10.6%) | 88.7% |
|  | 3 (CSSB) / ≥ 3 (TwinsUK) | 144 (5.7%) | 76.4% | 62 (4.6%) | 90.3% |
|  | 4 (CSSB only) | 92 (3.6%) | 79.3% |  |  |

Self-perceived COVID-19 recovery rates were 69% and 81% in CSSB & TwinsUK respectively, likely reflecting targeted recruitment of individuals with long COVID in CSSB. Work and Social Adjustment Scale collected for CSSB participants showed high levels of impairment of daily functioning among those who had not recovered from COVID-19 and who self-identified as having or had been diagnosed with long COVID (Figure S3). A larger proportion of COVID-19 cases were confirmed by self-reported positive antibody or antigen tests (versus suspected or based on medical advice) in CSSB vs. TwinsUK (CSSB: 87%, TwinsUK: 71%). At the time of reporting COVID-19 recovery status, most individuals were over a year, and a large proportion over two years, since the start of their COVID-19 infection (in CSSB: median: 687 days [IQR: 260, 898]; in TwinsUK: median: 411 days [IQR: 146, 755]). Just over half of COVID-19 cases dated from before the UK vaccination program commencing in December 2020 (CSSB: 52%, TwinsUK: 51%).

In composite measures of socio-demographic advantage and disadvantage, CSSB participants had proportionally more advantage and less disadvantage than TwinsUK participants.

A small number of CSSB participants were also members of TwinsUK (34 of 2,548, 1.3%), but it was not possible to determine whether these individuals were part of the TwinsUK analysis sample.

### Association between recovery and number of pre-pandemic social advantages or disadvantages

Associations were observed between recovery from COVID-19 and composite variables measuring socio-demographic advantage and disadvantage in both CSSB and TwinsUK cohorts (Figure 2, tabulated in Table S3) Individuals with a higher number of indicators of social disadvantage were less likely to recover (CSSB: OR = 0.74, 95% CI: 0.62-0.88, TwinsUK: OR = 0.79, 95% CI: 0.64-0.98 per indicator of disadvantage [of 5]), while individuals with more social advantage were more likely to recover (CSSB: OR = 1.26, 95% CI: 1.08-1.47, TwinsUK: OR = 1.36, 95% CI: 1.09-1.70 per indicator of advantage [of 4]). Inverse probability weights were used to adjust for response bias, and models adjusted for pre-pandemic health factors, which themselves were tested as exposures in secondary analyses (Figure S4). Models treating composite variables as categorical revealed the cumulative effect of multiple advantage or disadvantage to be approximately linear, in line with strong trends in crude recovery rates, which varied between approximately 52% to 80% with increasing advantage in CSSB and between 72% and 90% in TwinsUK (Figure S5). Associations persisted in models that included COVID-19 illness characteristics as potential mediators, which were themselves found to be strongly associated with recovery (Figure S6), with only weak, partial mediation with the addition of retrospectively reported symptom duration. Associations were also found in both males and females in sensitivity analyses where the CSSB sample was stratified by sex and sex was not included in the composite variables (Figure S7).

**Figure 2.**
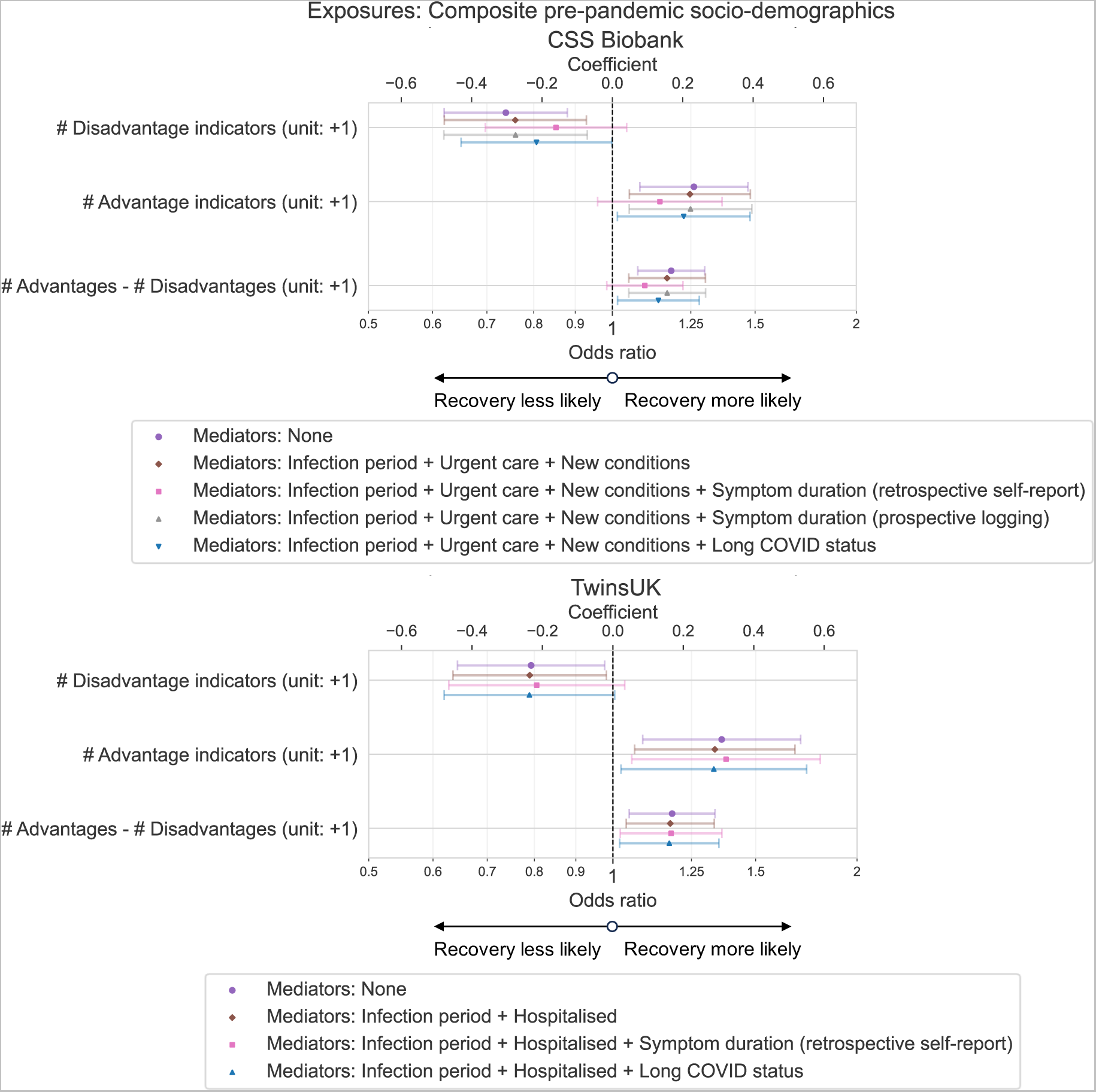
Associations between pre-pandemic indicators of socio-demographic advantage and/or disadvantage and recovery from COVID-19 in CSS Biobank and TwinsUK cohorts. Odds ratio and 95% confidence intervals from logistic regression models testing association between recovery from COVID-19 and composite measures of pre-pandemic of social advantage and disadvantage, among individuals with self-reported COVID-19 infection. Composite indicators were generated from sex, ethnic group, highest educational qualification, local area deprivation and pre-pandemic employment status. Results are shown for models including various COVID-19 illness characteristics as potential mediating factors. Models for both cohorts adjusted for age group, region, pre-pandemic self-rated general health, BMI, frailty and number of physical health conditions. TwinsUK models additionally adjusted for number of mental health conditions. Models weighted for inverse probability of participation.

### Effects of pre-pandemic socio-demographics

Associations with recovery were stronger for composite variables of socio-demographic advantage and/or disadvantage in comparison to the individual socio-demographics used to create composites (Figure 3). Individual socio-demographic associations (in terms of effect size and confidence levels) consistent across both cohorts were: lower likelihood of recovery for individuals with less than a university degree level qualification vs. degree level (CSSB: OR = 0.54, 95% CI: 0.39-0.76, TwinsUK: OR = 0.69, 95% CI: 0.48-1.01), individuals living in East Midlands region vs. London (CSSB: OR = 0.35, 95% CI: 0.19-0.65, TwinsUK: OR = 0.46, 95% CI: 0.22-0.98). Consistent direction of effect across cohorts were also seen for sex, ethnic group, and local area deprivation, but with 95% confidence intervals crossing the null in one or both cohorts. In CSSB we observed increased odds of recovery in individuals aged over 70 years; this was not observed in TwinsUK.

**Figure 3.**
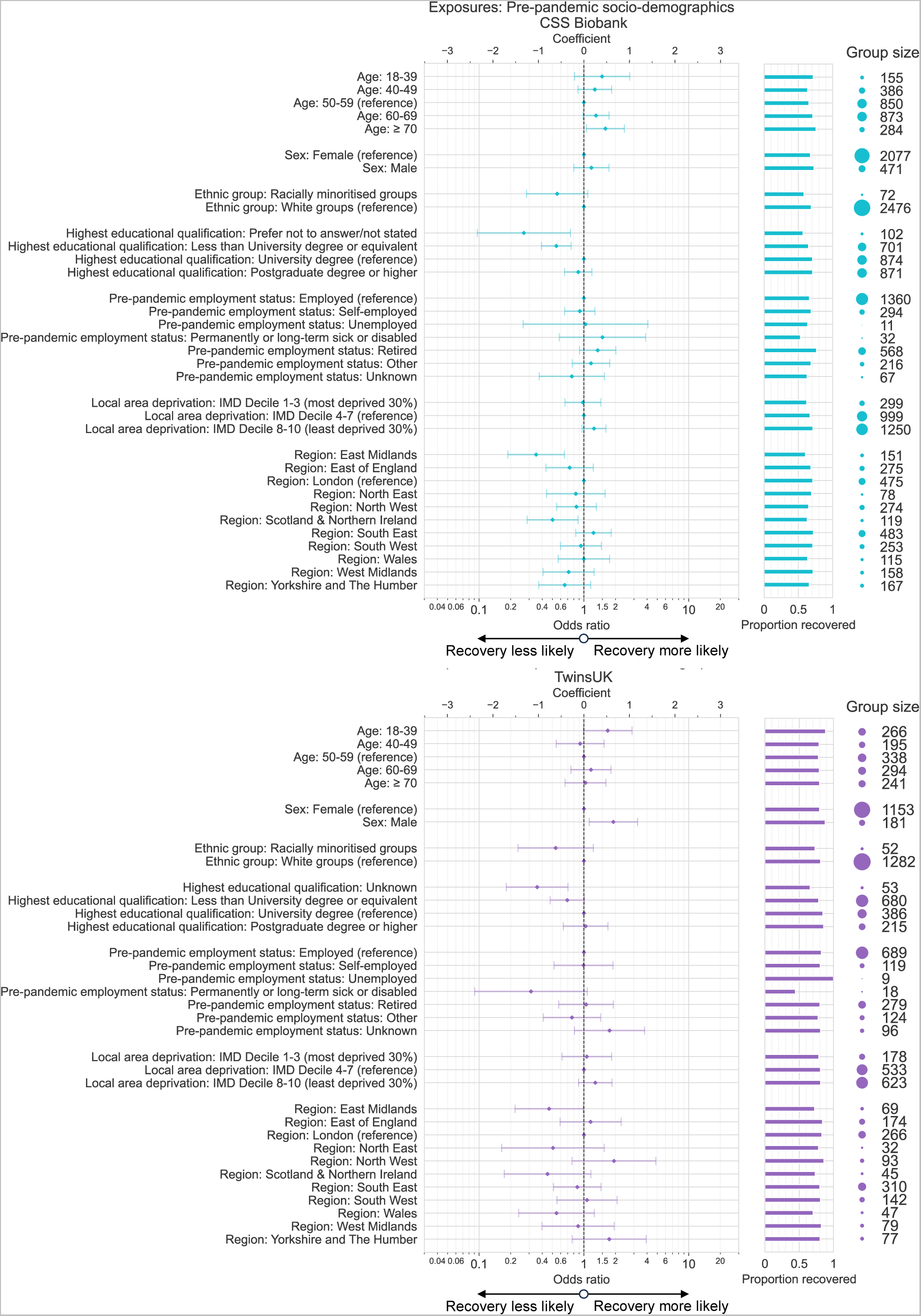
Associations between pre-pandemic socio-demographics and recovery from COVID-19 in CSS Biobank and TwinsUK cohorts. Odds ratio and 95% confidence intervals from logistic regression models testing association between recovery from COVID-19 and various pre-pandemic socio-demographic exposure variables, among individuals with self-reported COVID-19 infection. Results for each exposure variable originate from distinct models, including age, sex, ethnic group, education, pre-pandemic health characteristics and other pre-pandemic socio-demographic factors as potential confounding factors. Models weighted for inverse probability of participation.

Additional variables unique to CSSB and TwinsUK cohorts showed further associations (Figure S8, Table S1, Table S2), with higher likelihood of recovery for CSSB participants with first languages other than English (for whom, 81% were Western European languages), and lower likelihood of recovery for TwinsUK participants living in homes with damp, mould or vermin at the start of the pandemic.

### Associations with socio-demographics during the pandemic

Further associations were observed for socio-demographic factors reflecting status or experiences during the pandemic related to housing, employment, finances, access to health & social care and personal life (Figure 4, Table S1, Table S2). For such factors, directionality of causation was ambiguous due to unknown temporality of COVID-19 illness relative to reported experiences.

**Figure 4.**
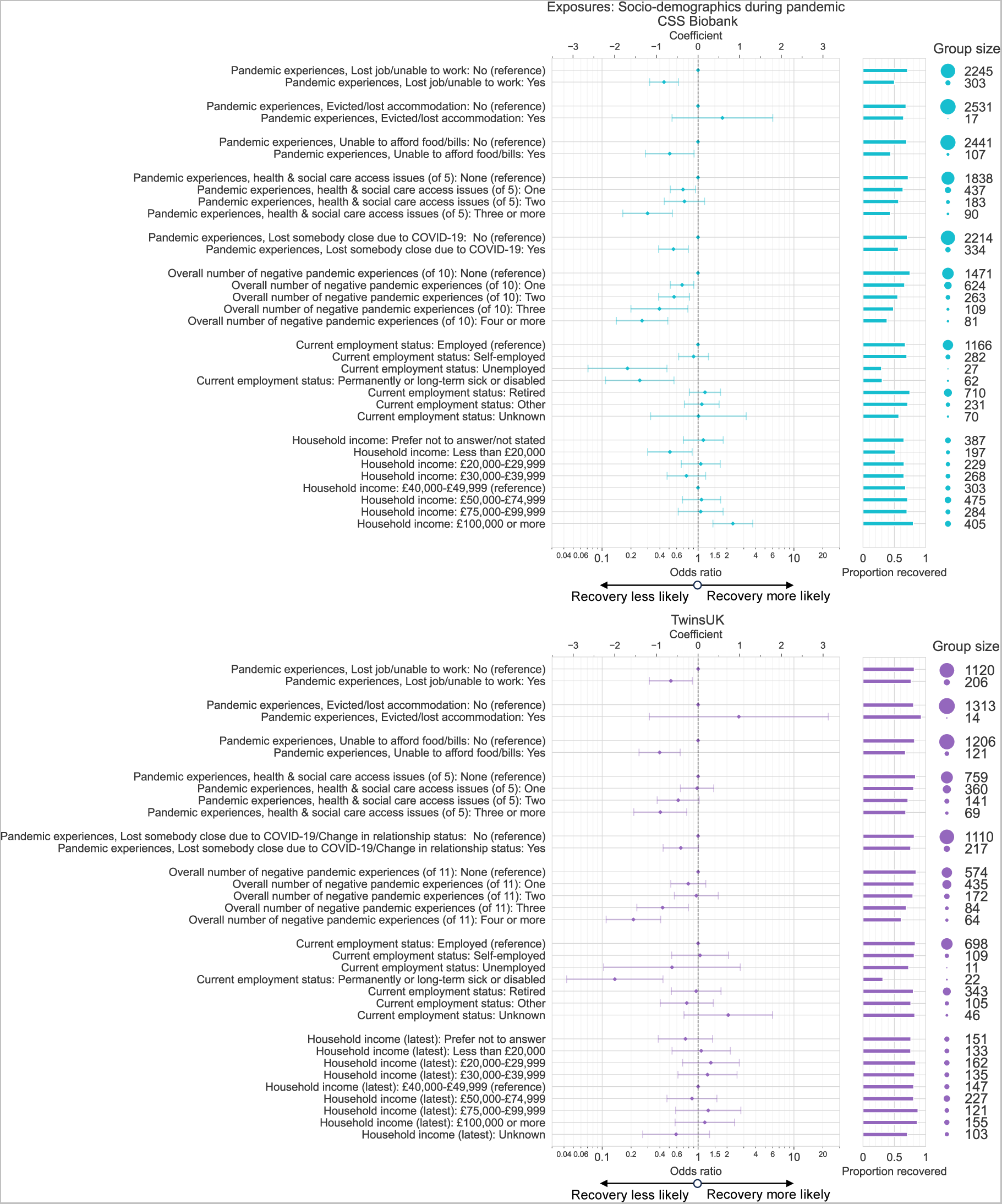
Associations between socio-demographics collected during the COVID-19 pandemic and recovery from COVID-19 in CSS Biobank and TwinsUK cohorts. Odds ratio and 95% confidence intervals from logistic regression models testing association between recovery from COVID-19 and various pre-pandemic socio-demographic exposure variables, among individuals with self-reported COVID-19 infection. Results for each exposure variable originate from distinct models, including age, sex, ethnic group, education, pre-pandemic socio-demographic factors and pre-pandemic health characteristics as potential confounding factors. Models weighted for inverse probability of participation.

Associations (in terms of effect size and confidence levels) consistent across both cohorts were: lower likelihood of recovery for individuals who lost their job or were unable to work (CSSB: OR = 0.44, 95% CI: 0.31-0.62, TwinsUK: OR = 0.52, 95% CI: 0.31-0.87), were unable to afford food or bills (CSSB: OR = 0.51, 95% CI: 0.28-0.91, TwinsUK: OR = 0.40, 95% CI: 0.24-0.65), experienced 3 or more types of health & social care access issues (from access to medication, community health services, community social care services, in/outpatient services, or mental health services) (CSSB: OR = 0.30, 95% CI: 0.16-0.54, TwinsUK: OR = 0.40, 95% CI: 0.21-0.76), lost somebody close due to COVID-19 (CSSB: OR = 0.55, 95% CI: 0.39-0.79, TwinsUK: OR = 0.66, 95% CI: 0.43-1.01). A composite variable counting the overall number of negative experiences showed stronger associations than the individual experiences. Current employment status of permanently or long term sick or disabled vs. employed was associated with lower likelihood of recovery vs. employed in both cohorts. A trend of increasing likelihood of recovery with increasing current household income observed in CSSB was not seen in TwinsUK.

Additional variables unique to TwinsUK cohorts showed further associations (Figure S8), with lower likelihood of recovery for TwinsUK participants who experienced significant stress in the early months of the pandemic, or made new claims for government credit/benefits.

### Do pandemic experiences explain the social disparity in recovery?

Finally, we tested whether social disparities in recovery following COVID-19 (Figure 2) were mediated by differences in adverse employment, financial, health & social care access, and personal experiences during the COVID-19 pandemic, which were themselves found to be strongly associated with recovery (Figure 4). We found partial weak mediation by adverse experiences in both cohorts (Figure 5). Individual adverse experiences showed marginal and inconsistent changes in effect sizes, with the largest moderation of effect size seen in both cohorts for the overall count of adverse experiences across all domains. However, these experiences did not fully explain the observed social disparity in recovery.

**Figure 5.**
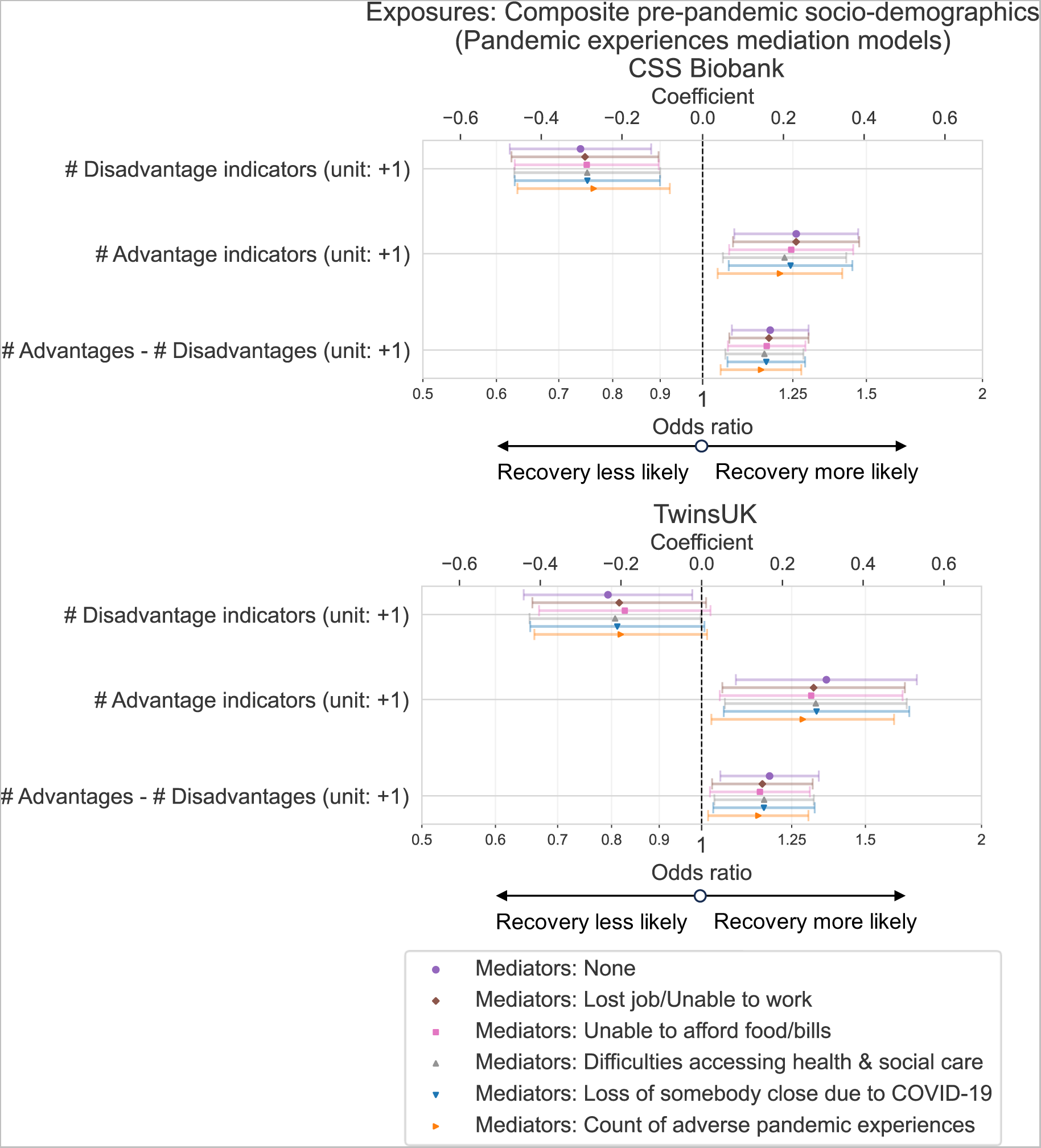
Testing mediation of social gradients in recovery from COVID-19 by adverse pandemic experiences in CSS Biobank and TwinsUK cohorts. Odds ratio and 95% confidence intervals from logistic regression models testing association between recovery from COVID-19 and composite measures of pre-pandemic of social advantage and disadvantage, among individuals with self-reported COVID-19 infection. Composite indicators were generated from sex, ethnic group, highest educational qualification, local area deprivation and pre-pandemic employment status. Results are shown for models including various adverse pandemic experiences as potential mediating factors. Models for both cohorts adjusted for age group, region, pre-pandemic self-rated general health, BMI, frailty and number of physical health conditions. TwinsUK models additionally adjusted for number of mental health conditions. Models weighted for inverse probability of participation.

## Discussion

### Key points

We found strong relationships between recovery from COVID-19 and measures of socio-demographic advantage and disadvantage in two different UK longitudinal population studies, COVID Symptom Study Biobank (CSSB) and TwinsUK. Holding multiple advantageous statuses/positions (based on existing systems of power) across sex, ethnic group, education, local deprivation and pre-pandemic employment status led to a higher likelihood of reporting full recovery from COVID-19, in models that accounted for age, region of residence and pre-pandemic health factors (Figure 2). In contrast, multiple disadvantages prior to the pandemic were associated with a lower likelihood of recovery and higher rates of ongoing symptoms, on average more than a year since infection. Composite socio-demographic variables were stronger predictors of COVID-19 recovery than the individual socio-demographic variables from which they were generated, showing the importance of considering social determinants in combination rather than in isolation.

Social gradients in recovery rate were not explained by differences in the timing and severity of COVID-19 illness along socio-demographic lines, with evidence of weak partial mediation only, and associations were seen in both female and male sex in sex-stratified models (Figure S7). Non-recovery was also associated with living in housing with mould, damp or vermin issues at the start of the pandemic, as well as adverse employment, financial, health & social care access, and personal relationship experiences during the pandemic. We again found that social gradients in recovery were not fully explained by differences in these experiences along socio-demographic lines, with evidence of weak partial mediation only.

### Interpretation

With associations not explained by extensive adjustment for pre-existing health, and not explained by difference in COVID-19 illness factors or adverse experiences during the pandemic, our results raise the question: how does our observed social inequality in recovery from COVID-19 operate?

The observed inequality could be theorised as a consequence of structural differences in opportunity and value assigned to certain groups [27], and/or in terms of differences between groups in social, cultural and economic capital [43]. Differences in such resources and experiences may affect recovery through the many pathways identified in biopsychosocial models linking socioeconomic status and health more generally [44]. Although we found that adverse pandemic experiences did not fully mediate disparities in COVID-19 recovery, previous reports have found associations between long COVID and (in)adequate income, sickness-related absence from work and economic (in)activity [11,45]. These reports illustrate how ongoing symptoms following COVID-19 may feed into pre-existing negative cycles between low economic capital/poverty and ill health [46].

We note strong associations between non-recovery and experiences of being unable to access health & social care, which in principle should be freely available to all within the UK National Health Service. This In previous other reports. This is consistent with reports finding that health care for COVID-19 has been most accessible for the most structurally advantaged. Gendered and racialised health care experiences were described by individuals living with long COVID in Bradford, UK [47], while higher rates of long COVID coding in electronic healthcare records have been found for white ethnic groups and those living in less deprived areas (in conflict with prevalence estimates) [48,49]. Other pathways/mechanisms that could be explored in further work include: access to social support networks; inequalities in resources, conditions or responsibilities that affect recuperation as previously identified [50–52]; and biological stressors such as adverse experiences, including sexism and racism [53–55]. Additionally, unmeasured biological confounding such as genetic differences may go further towards explaining the observed social gradient. Equally, there may be more complex interactions between COVID-19 illness and pre-pandemic health than our models accounted for.

To put our results into context, socio-demographic factors including income, isolation, social support, race/ethnicity, education and local area deprivation have also been found to be social determinants of recovery from mental illness [56,57], poor general physical health [58], and following hospitalisation in critical care [59]. Such results suggest that social factors likely play a significant role in recovery from illness in general, not only in the case of COVID-19.

### Limitations

We note the limitations in our study. CSSB cohort recruitment was conditioned on the use of a smartphone app, in addition to self-reported SARS-CoV-2 infection status and COVID-19 symptom duration. However comparable associations were found in TwinsUK, where participation was not conditioned upon COVID-19 illness. Compared to the general United Kingdom population, both TwinsUK and CSSB cohorts are overrepresented by white ethnic groups, female sex and those living in more affluent areas. Both cohorts also rely on voluntary participation. As such, there is potential for collider bias to operate [60], which we attempted to address by using inverse probability weighting in models.

Analysis of certain groups known to be subject to structural social disadvantage was not possible due to absence of data or limited by sample size, such as for distinct racially minoritised ethnic groups that were undesirably aggregated into a single group. For the same reasons, we did not test for intersectional effects (smaller/larger than individual additive effects) as in other reports [61–63], and generated *non-specific* measures of the *overall number* of indicators of social disadvantage and/or advantage. In these measures, each indicator was treated as having equal weight, despite differences in group size and strength of associations observed when tested individually. The choice of socio-demographic variables and indicators of social (dis)advantage in our intersectional analyses were subjective and some were UK-specific metrics, although based on known systems of power and social inequalities in health. Variable choice was also limited by pre-pandemic data available across CSSB and TwinsUK cohorts, although associations were found to be robust to modifications in sensitivity analyses.

### Summary

In summary, our analyses suggest a striking social health gradient in self-perceived recovery from ongoing symptoms following COVID-19 infection, which correlates strongly with functional impairment. COVID-19 may be a model illness for demonstrating social inequalities in recovery more generally. In turn, disparities in recovery may accumulate and contribute substantially to broader social inequalities in health. Targeted support for individuals during recovery periods from acute illness may help to address inequalities, informed by further investigation of the biopsychosocial mechanisms that underly social inequalities in recovery following COVID-19 as well as other illnesses.

## Contributors

Following CRediT framework: https://www.elsevier.com/authors/policies-and-guidelines/credit-author-statement.

Conceptualisation: NJC, CJS, with contribution from all authors

Funding acquisition: CJS, ELD

Project administration: VB, MPG, CJS

Methodology: NJC, CJS

Formal analysis: NJC

Investigation: VB, NJC

Visualisation: NJC

Data curation: VB, LSC, NJC, EK, BM, CHS

Writing – original draft: NJC, CJS, JDC, RCEB, VB, MPG

Writing – review and editing: All authors

## Data sharing

For the purposes of open access, the author has applied a Creative Commons Attribution (CC BY) licence to any Accepted Author Manuscript version arising from this submission.

Access to data in the CSS Biobank is available to bona fide health researchers on application to the CSS Biobank Management Group. Further details are available online at https://cssbiobank.com/information-for-researchers including application forms and contact information. Analysis code used in this study is available openly on GitHub at https://github.com/nathan-cheetham/CSSBiobank_COVIDRecovery. Anonymised COVID Symptom Study data are available to researchers to be shared with researchers according to their protocols in the public interest through Health Data Research UK (HDRUK) and Secure Anonymised Information Linkage consortium, housed in the UK Secure Research Platform (Swansea, UK) at https://web.www.healthdatagateway.org/dataset/fddcb382-3051-4394-8436-b92295f14259.

## Ethics statement

Yorkshire & Humber NHS Research Ethics Committee gave ethical approval for the COVID Symptom Study Biobank, Ref: 20/YH/0298.

All waves of TwinsUK have received ethical approval associated with TwinsUK Biobank (19/NW/0187), TwinsUK (EC04/015) or Healthy Ageing Twin Study (H.A.T.S) (07/H0802/84) studies from HRA/NHS Research Ethics Committees.

## Supporting information

STROBE guidelines

ICMJE disclosure forms

## Data Availability

For the purposes of open access, the author has applied a Creative Commons Attribution (CC BY) licence to any Accepted Author Manuscript version arising from this submission.
Access to data in the CSS Biobank is available to bona fide health researchers on application to the CSS Biobank Management Group. Further details are available online at https://cssbiobank.com/information-for-researchers including application forms and contact information. Analysis code used in this study is available openly on GitHub at https://github.com/nathan-cheetham/CSSBiobank_COVIDRecovery. Anonymised COVID Symptom Study data are available to researchers to be shared with researchers according to their protocols in the public interest through Health Data Research UK (HDRUK) and Secure Anonymised Information Linkage consortium, housed in the UK Secure Research Platform (Swansea, UK) at https://web.www.healthdatagateway.org/dataset/fddcb382-3051-4394- 8436-b92295f14259.

## Acknowledgements

We thank COVID Symptom Study Biobank participants, in particular those who participated in various studies while experiencing ongoing symptoms. We are grateful to the CSS Biobank Volunteer Advisory Panel for their input on the development of the biobank, and feedback on the design of this study and its significance for patients and the wider public.

We are grateful to staff at Zoe Ltd (including Christina Hu and Joan Capdevila Pujol) for their work on the CSS app, for enabling recruitment to the CSS Biobank and sharing and maintaining CSS data. We are grateful to data and program team staff at TwinsUK (including Andrew Anastasiou, Genevieve Lachance, Darioush Yarand) for work on CSSB recruitment, setup and data management. We thank Katie Doores, Michael Malim, Carl Graham, Jeffrey Seow, Sam Acors, and Neo Kouphou for antibody testing of participant blood samples.

The CSS Biobank is supported by the Chronic Disease Research Foundation. TwinsUK is funded by the Medical Research Council (MRC), Wellcome LEAP, Wellcome Trust, Engineering & Physical Sciences Research Council, Biotechnology and Biological Sciences Research Council, Versus Arthritis, European Commission, Chronic Disease Research Foundation (CDRF), Zoe Ltd, the National Institute for Health and Care Research (NIHR) Clinical Research Network (CRN) and Biomedical Research Centre based at Guy’s and St Thomas’ NHS Foundation Trust in partnership with King’s College London.

NJC, EJT, CJS and JDC were supported by the NIHR CONVALESCENCE grant [COV-LT-0009]. RSP is a fellow on the Multimorbidity Doctoral Training Programme for Health Professionals, which is supported by the Wellcome Trust [223499/Z/21/Z]. EM was supported by MRC [MR/R016372/1] and NIHR [NIHR134293]. LSC is supported by Wellcome Trust grant [215010/Z/18/Z].

Authors affiliated with King’s College London are also supported by the Wellcome Trust / Engineering and Physical Sciences Research Council Centre for Medical Engineering at King’s College London (KCL, [203148/Z/16/Z]) and the UK Department of Health via the NIHR comprehensive Biomedical Research Centre award to Guy’s & St Thomas’ NHS Foundation Trust (GSTT) in partnership with KCL and King’s College Hospital NHS Foundation Trust. ZOE Ltd provided in-kind support for all aspects of building, running and supporting the COVID Symptom Study app and service to all users worldwide.

## The COVID Symptom Study Biobank Consortium

Michela Antonelli^1^, Vicky Bowyer^2^, Julia Brown^2,3^, Liane Canas^1^, Joan Capdevila Pujol^4^, Nathan Cheetham^2^, Lynn Cherkas^2^, Jie Deng^1^, Katie Doores^5^, Emma Duncan^2,6^, Maria Paz Garcia^2^, Alexander Hammers^1,7^, Deborah Hart^2^, Nicholas Harvey^2^, Adrian Hopper^8^, Christina Hu^4^, Eric Kerfoot^1^, Michael Malim^5^, Marc Modat^1^, Erika Molteni^1^, Benjamin Murray^1^, Ayrun Nessa^2^, Sebastien Ourselin^1^, Tim Spector^2^, Claire Steves^2,9^, Carole Sudre^1,10,11^, Samuel Wadge^2^, Jonathan Wolf^4^

^1^School of Biomedical Engineering & Imaging Sciences, King’s College London, London, United Kingdom.

^2^Department of Twin Research and Genetic Epidemiology, King’s College London, London, United Kingdom.

^3^Medical Student, James Cook University, Australia.

^4^ Zoe Ltd, 164 Westminster Bridge Road, London, United Kingdom.

^5^Department of Infectious Diseases, King’s College London, London, United Kingdom.

^6^Department of Endocrinology, Guy’s and St Thomas’ NHS Foundation trust, London, United Kingdom.

^7^King’s College London & Guy’s and St Thomas’ PET Centre, King’s College London, London, United Kingdom.

^8^Guy’s and St Thomas’ NHS Foundation trust, London, United Kingdom.

^9^Department of Ageing and Health, Guy’s and St Thomas’ NHS Foundation trust, London, United Kingdom.

^10^MRC Unit for Lifelong Health and Ageing, Department of Population Science and Experimental Medicine, University College London, London, United Kingdom.

^11^Centre for Medical Image Computing, Department of Computer Science, University College London, London, United Kingdom.

