## Supplementary information for "Social determinants of recovery from ongoing symptoms following COVID-19 in two UK longitudinal studies: a prospective cohort study"

#### Authors & affiliations

Nathan J. Cheetham[1]*, Vicky Bowyer[1], María Paz García[1], Ruth C. E. Bowyer[1,2], J. D. Carpentieri[3], Andy Guise[4], Ellen J. Thompson[1,5], Carole H. Sudre[6,7,8], Erika Molteni[8], Michela Antonelli[8], Rose S. Penfold[9], Nicholas R. Harvey[1], Liane S. Canas[8], Khaled Rjoob[6], Benjamin Murray[8], Eric Kerfoot[8], The COVID Symptom Study Biobank Consortium[^], Alexander Hammers[8,10], Emma L. Duncan[1,11], Claire J. Steves[1,11]*

* Corresponding authors

1 Department of Twin Research and Genetic Epidemiology, King’s College London, London, United Kingdom

2 The Alan Turing Institute, London, United Kingdom

3 Institute of Education, University College London, London, United Kingdom

4 Department of Population Health Sciences, King’s College London, London, United Kingdom

5 School of Psychology, University of Sussex, Falmer, United Kingdom

6 MRC Unit for Lifelong Health and Ageing, Department of Population Science and Experimental Medicine, University College London, London, United Kingdom

7 Centre for Medical Image Computing, Department of Computer Science, University College London, London, United Kingdom

8 School of Biomedical Engineering & Imaging Sciences, King’s College London, London, UK

9 Edinburgh Delirium Research Group, Ageing and Health, Usher Institute, University of Edinburgh, Edinburgh, United Kingdom

10 King's College London & Guy's and St Thomas' PET Centre, King's College London, London, United Kingdom

11 Guy’s & St Thomas’s NHS Foundation Trust, London, United Kingdom

^A list of authors and their affiliations appears below

#### The COVID Symptom Study Biobank Consortium

Michela Antonelli[1], Vicky Bowyer[2], Julia Brown[2,3], Liane Canas[1], Joan Capdevila Pujol[4], Nathan Cheetham[2], Lynn Cherkas[2], Jie Deng[1], Katie Doores[5], Emma Duncan[2,6], Maria Paz Garcia[2], Alexander Hammers[1,7], Deborah Hart[2], Nicholas Harvey[2], Adrian Hopper[8], Christina Hu[4], Eric Kerfoot[1], Michael Malim[5], Marc Modat[1], Erika Molteni[1], Benjamin Murray[1], Ayrun Nessa[2], Sebastien Ourselin[1], Tim Spector[2], Claire Steves[2,9], Carole Sudre[1,10,11], Samuel Wadge[2], Jonathan Wolf[4]

1 School of Biomedical Engineering & Imaging Sciences, King’s College London, London, United Kingdom.

2 Department of Twin Research and Genetic Epidemiology, King’s College London, London, United Kingdom.

3 Medical Student, James Cook University, Australia.

4 Zoe Ltd, 164 Westminster Bridge Road, London, United Kingdom.

5 Department of Infectious Diseases, King’s College London, London, United Kingdom.

6 Department of Endocrinology, Guy’s and St Thomas’ NHS Foundation trust, London, United Kingdom.

7 King's College London & Guy's and St Thomas' PET Centre, King's College London, London, United Kingdom.

8 Guy’s and St Thomas’ NHS Foundation trust, London, United Kingdom.

9 Department of Ageing and Health, Guy’s and St Thomas’ NHS Foundation trust, London, United Kingdom.

10 MRC Unit for Lifelong Health and Ageing, Department of Population Science and Experimental Medicine, University College London, London, United Kingdom.

11 Centre for Medical Image Computing, Department of Computer Science, University College London, London, United Kingdom.

#### Data collection timeline

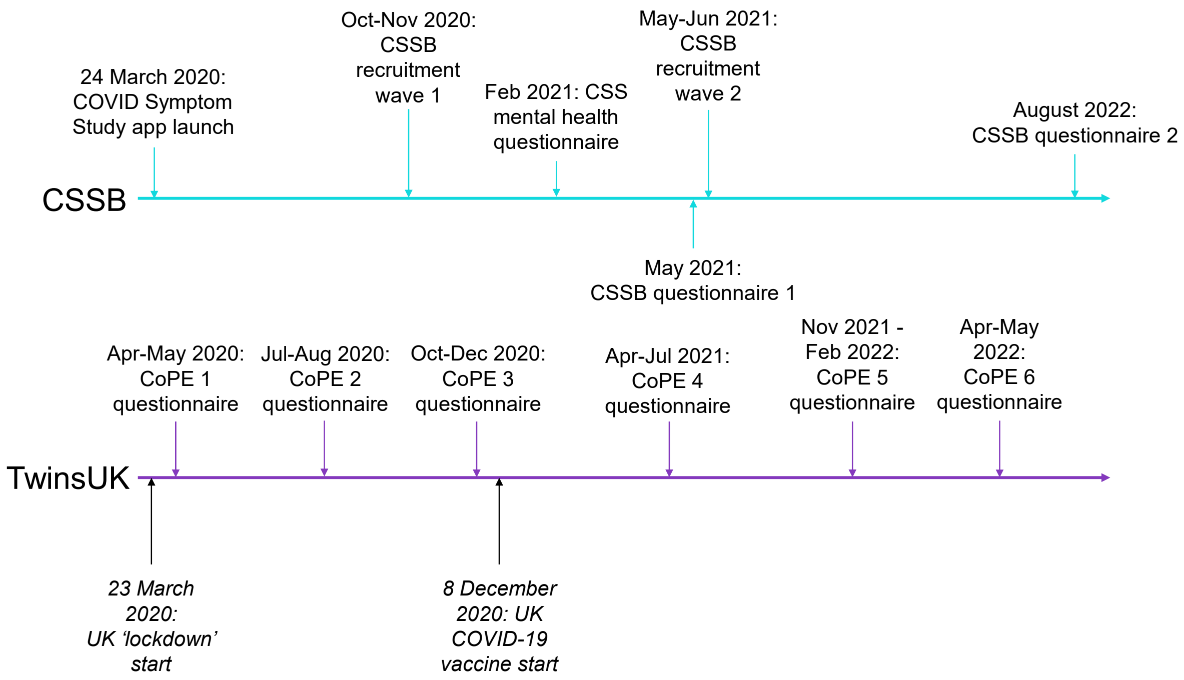

Figure S 1. **Timeline of data collections for CSSB and TwinsUK.** CSS: COVID Symptom Study app, CSSB: COVID Symptom Study Biobank.

#### 1. CSSB Data collection

##### *Socio-demographic characteristics*

Information on age group at the time of the 2022 CSSB questionnaire was derived from date of birth self-reported at CSSB consent (2022 – Year of Birth).

Information on sex assigned at birth (female/intersex/male/prefer not to say) was self-reported at registration to the CSS app.

Information on race/ethnicity was collected at CSSB consent with the question "what is your ethnic group?", using UK 2021 census categories [1]. Due to small number of individuals in certain individual groups, responses were grouped into White groups (including all White groups), and all other response options (including all Asian/Asian British, Black/Black British/Caribbean/African, Mixed/Multiple ethnic groups, and Other ethnic groups), referred to collectively as Racially Minoritised groups following recent recommendations [2].

First language was self-reported in the August, 2022 CSSB questionnaire, and grouped into English and Other due to small number of individuals with first languages other than English.

Information on highest educational qualification was collected in the August, 2022 CSSB questionnaire "What is the highest academic/educational qualification (or its nearest equivalent) you have received?". The category "Less than University degree or equivalent" comprised "Did not complete secondary school" (approximately < 12 years schooling), "General Certificate of Secondary Education (GCSE) or (General National Vocational Qualification) GNVQ or equivalent" (~12 years schooling), and "A-Levels or advanced GNVQ or equivalent" (~14 years schooling) options. "Postgraduate degree or higher" comprised "Postgraduate degree or higher" and "PhD" options.

Local area deprivation and UK geographic region of residence were derived from address data collected at CSSB consent. Deprivation was measured by the Index of Multiple Deprivation (IMD) quintiles of lower super output area rank, with data from England 2019 [3], Wales 2019 [4], Scotland 2020 [5], and Northern Ireland 2017 [6]. Individuals living in Scotland and Northern Ireland were grouped together in "Scotland/Northern Ireland" category, due to small number of participants residing in Northern Ireland. Scotland was chosen as the nearest other UK region.

Equivalent data for education level and residential address immediately prior to the COVID-19 pandemic was not available and responses were assumed to represent pre-pandemic statuses.

Information on pre-pandemic employment status was self-reported in the May, 2021 CSSB questionnaire in the question "Which one of these best describes what you were doing before the COVID-19 pandemic? If you were doing more than one activity, please choose the activity you spent the most time doing". Due to small numbers of individuals, "Other" was a heterogeneous group comprising "In unpaid/voluntary work", "In education at school/college/university, or in an apprenticeship", "Looking after home or family", "Other" options.

Current employment status was self-reported in the August, 2022 CSSB questionnaire, with "other" options grouped as for the pre-pandemic equivalent.

Current yearly gross household income was self-reported in the August, 2022 CSSB questionnaire with the question "What is the total yearly income before tax received by your household? This includes all those who can earn and live in the same household as yourself."

Adverse experiences during the pandemic were measured in the August, 2022 CSSB questionnaire with the question: "Have you experienced any of the following during the COVID-19 pandemic?" (Yes/No/Prefer not to say). Adverse employment, housing and personal experiences were described by options "Lost your job/been unable to do paid work", "Evicted/lost accommodation", and "You lost somebody close to you due to COVID-19" respectively. Adverse financial experiences were described by "Unable to pay bills" and "Unable to afford food", which were combined in analyses. The number of general health and social care access issues experienced during the pandemic was generated from the number of Yes responses to the 5 response options: "Unable to access required medication”, “Unable to access health services in the community, for instance, GP/community physiotherapy/nurse/podiatrist/dentist”, “Unable to access the community social care services or voluntary sector support you need, for instance, from carers or day centres”, “Unable to access inpatient or outpatient appointments booked at a hospital for a consultation, investigation, treatment, or surgery”, and “Unable to access appointment for cognitive behaviour therapy, counselling, or psychological therapy”.

##### *Health characteristics*

Pre-pandemic general health was self-reported in the May, 2021 CSSB questionnaire with the question “In general, in the 3 months before the COVID-19 outbreak in March 2020, would you say your health was...”.

Body mass index (BMI) was derived from self-reported height and weight collected at CSSB consent.

Frailty, a measure of age-related decline in physiological reserve and function [7], was measured using the PRISMA-7 scale [8], collected at registration with the CSS app.

Number of physical health conditions was measured from 6 self-reported conditions (asthma, cancer, diabetes, heart disease, lung disease, kidney disease) collected at registration for the CSS app.

Number of mental health conditions was measured from self-reported diagnoses of 16 conditions (Generalised anxiety order; Panic disorder; Specific phobias; Obsessive compulsive disorder; Post-traumatic stress disorder; Social anxiety disorder; Agoraphobia; Depression; Attention deficit or attention deficit and hyperactivity disorder; Autism, Asperger's or autistic spectrum disorder; Eating disorder (e.g. bulimia nervosa; anorexia nervosa; psychological over-eating or binge-eating), Personality disorder; Mania, hypomania, bipolar or manic depression; Schizophrenia; Substance use disorder; Any other type of psychosis or psychotic illness) collected in a February, 2021 CSS questionnaire.

Equivalent data used to derive BMI, frailty and number of physical health conditions representing status immediately prior to the COVID-19 pandemic was not available and responses were assumed to represent pre-pandemic statuses.

Work and social adjustment scale (WSAS) [9], collected in the August, 2022 CSSB questionnaire, asked individuals to assess how their health impaired their ability to do day-to-day tasks over the last 3 months.

##### *COVID-19 illness characteristics*

COVID-19 infection history was measured with the question “How many times do you think you have ever had COVID-19? Please include now if you think you currently have COVID-19 symptoms but have not confirmed it.”. For each reported infection, participants were asked what evidence supported their infection ("How do/did you know you had it (COVID-19)?"), the start date of the infection/illness ("When do you think you had COVID-19? Please use your best estimate if you can't remember the exact date."), the duration of COVID-19 symptoms ("How long did you have or have you had continuous symptoms?") and the duration for which they were not able to function as normal ("How long were you, or have you been unable to function as normal due to COVID-19 symptoms?"). Where multiple infections were reported, information was obtained for the single infection with the longest reported symptom duration, then earliest date of infection start, then strongest evidence of infection.

In addition to retrospective self-reported symptom duration, prospective symptom reporting was available for CSSB participants via self-reporting using the CSS app. Symptom duration from prospective symptom reporting in the app was estimated using methods described in previous reports [10].

Infection period was derived from self-reported date of infection start. Periods were defined based on changes in dominant SARS-CoV-2 variant from COG-UK Mutation Explorer data available at https://sars2.cvr.gla.ac.uk/cog-uk/.

Whether individual accessed urgent care during their COVID-19 illness was assessed from the questions "What type of medical help did you access WITHIN the first 4 weeks of (/MORE THAN 4 weeks after) the start of your symptoms that you think may have been caused by COVID-19?", from selection of options "Visited A&E or walk-in centre" or "Called an ambulance".

Number of new conditions due to COVID-19 was generated from responses to the question "Please tell us which new health condition, illness, or disability you have been told you have since March 2020.", from 22 health condition options, where individuals selected Yes to "Has a doctor told you that this new health condition developed because of COVID-19 infection?".

Self-reported or diagnosed long COVID was measured by the question "Have you ever received a diagnosis of long COVID or post-COVID syndrome?", with "No, but I do believe I have or have had Long COVID" or "Yes" taken as indicative of long COVID and "No, and I do not believe I have or have had Long COVID" as not.

#### 2. TwinsUK Data collection

***Socio-demographic characteristics***

Information on sex assigned at birth and age group at time of latest questionnaire (2022 – Year of Birth) from date of birth was collected at TwinsUK registration. Information on race/ethnicity and highest educational qualification was collected as part of regular TwinsUK longitudinal questionnaires.

Local area deprivation and UK geographic region of residence were derived from most recent address data as of April 1, 2020.

Information on pre-pandemic employment status was self-reported in CoPE #1, #4 and #5 questionnaires, and current employment status collected in CoPE #4, #5 and #6, using the same question and response options as described for CSSB. Earliest and latest valid responses were taken for pre-pandemic and current employment status respectively.

Yearly gross household income was collected as part of TwinsUK longitudinal questionnaires. Pre-pandemic income was generated from the latest available response between January 1, 2017 and April 1, 2020, while latest income considered the latest available response after January 1, 2017.

Adverse experiences during the pandemic were measured in every CoPE #1 to #6, with the question: "Have you experienced any of the following as a result of COVID-19?". Response options were expanded over earlier CoPE rounds based on increased understanding of adverse experiences, to be equivalent to CSSB from CoPE #4 onwards. Responses in TwinsUK were treated as equivalent to CSSB, and interpreted as experiences during the COVID-19 pandemic in both cases, despite differences in question wording. For responses across multiple questionnaires, the maximal responses were taken.

Variables unique to TwinsUK:

Housing tenure and housing problems with damp, mould or vermin were self-reported in CoPE #1.

Credit or benefits claims prior to the pandemic was collected in CoPE #1 and #6 with the question "Before the pandemic (March 2020), did you or your partner regularly claim for the following?", with Yes/No for the following response options: Free school meals, Universal credit, Pension credit, Employment Support allowance, Statutory sick pay, Housing benefit, council tax benefit, carers allowance and Personal Independence Payment (PIP). New credit or benefits claims during the pandemic was collected in CoPE #2, #3 and #6 with the question "Since the start of the pandemic (March 2020), have you or your partner ever made any new claims for the following?", with the same 6 response options as the pre-pandemic question, plus the additional option "A grant through the new self-employment income support scheme" which described the UK government scheme to cover wages for those whose work was affected by the COVID pandemic. For responses across multiple questionnaires, the maximal responses were taken.

Number of significant stressors was derived from the following question asked in CoPE #1 and #2: “Have any of these things been causing you SIGNIFICANT stress? E.g. They have been constantly on your mind or have been keeping you awake at night”, counting the number of ‘Yes’ responses to the following 16 response options: Marriage or other romantic relationship, Friends or family living in your household, Friends or family living outside your household, Neighbours, Your pet(s), Work (even if you feel your job is safe), Losing your job/unemployment, Finances, Getting medication, Getting food, Your own safety/security, Internet access, Boredom, Future plans, Catching COVID-19, Becoming seriously ill from COVID-19. For responses across multiple questionnaires, the maximal responses were taken.

Number of caring responsibilities was derived from the following questions asked in CoPE #1, #2 and #3: “Are you currently responsible for the care or support of any of the following INSIDE the home?” and “Are you currently responsible for the care or support of any of the following OUTSIDE the home?”, counting the number of ‘Yes’ responses to the following response options which were asked in both questions: Elderly relatives or friends, People with long-term conditions or disabilities, Grandchildren, Children (under 18). For responses across multiple questionnaires, the maximal responses were taken.

***Health characteristics***

Pre-pandemic general health was self-reported in CoPE #1 and #2 questionnaires and was the same as described in CSSB. For responses across multiple questionnaires, the earliest valid response was taken.

Body mass index (BMI) was derived from latest available height and weight data prior to April 1, 2020, collected from either self-report in regular longitudinal questionnaires or clinic visits.

Frailty, measured using the PRISMA-7 scale, was collected in CoPE #1 and #2 questionnaires. For responses across multiple questionnaires, the earliest valid response was taken.

Number of physical health and mental health conditions were collected from latest available responses in regular longitudinal questionnaires. The same physical conditions as collected in CSSB were chosen for cross-cohort harmonisation. Mental health conditions available in TwinsUK were limited to 4 conditions: bipolar disorder, anxiety/stress disorder, depression and eating disorder.

***COVID-19 illness characteristics***

COVID-19 infection history was derived based on self-report in each of CoPE #1 to #6. As in CSSB, information was collected on evidence of infection, start date of infection/illness, duration of COVID-19 symptoms, and duration for which they were not able to function as normal. Where multiple infections were reported, information was obtained for the single infection with the longest reported symptom duration, then earliest date of infection start, then strongest evidence of infection.

Infection period was derived from self-reported date of infection start as in CSSB. COVID-19 illness severity was measured in TwinsUK through questions on whether individuals were admitted to hospital as a result of COVID-19, asked in CoPE #2 through #6. For responses across multiple questionnaires, the maximal responses were taken. Number of new conditions due to COVID-19 asked to CSSB participants was not available in TwinsUK.

Self-reported or diagnosed long COVID was measured using the same question as described in CSSB, asked in CoPE #5 and #6. For responses across multiple questionnaires, the latest valid response was taken. We note however, that the first iteration of the question in CoPE #5 had only "Yes" and "No" as response options, leaving individuals' self-perception of whether they had long COVID unspecified. Therefore, an additional category, "No (self-reported long COVID status unknown)" was included in TwinsUK analyses for responses based on this question.

#### Sample selection flow diagram

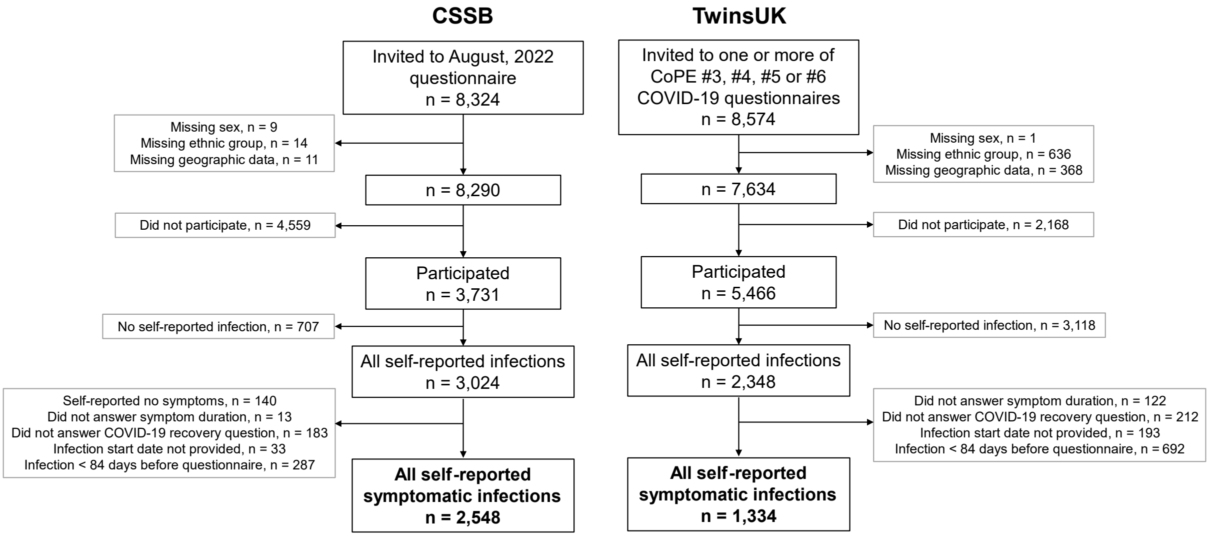

Figure S 2. **Sample selection flow diagram.** Exclusions are identified in grey boxes.

#### Extended sample characteristics & multivariable logistic regression results

##### *COVID Symptom Study Biobank*

Table S 1. **Extended sample characteristics and results of multivariable logistic regression models, CSSB cohort.** Odds ratios and confidence intervals (CI) are presented for multivariable logistic regression models testing association between recovery from COVID-19 and exposure of interest, after adjustment as appropriate from the hypothesised directed acyclic graph (DAG), and weighting for inverse probability of participation.

| | COVID Symptom Study Biobank (CSSB) | | | | | | |
|---|---|---|---|---|---|---|---|
| **Domain** | **Variable** | **Category** | **Group size, N (%)** | **COVID-19 recovery (%)** | **Odds ratio** | **95% CI (lower)** | **95% CI (upper)** |
| **Individual pre-pandemic demographics** | Age | 18-39 | 155 (6.1%) | 71.6% | 1.49 | 0.81 | 2.74 |
| | Age | 40-49 | 386 (15.1%) | 63.5% | 1.27 | 0.88 | 1.84 |
| | Age | 50-59 (reference) | 850 (33.4%) | 65.4% | 1.00 | | |
| | Age | 60-69 | 873 (34.3%) | 71.1% | 1.30 | 0.98 | 1.74 |
| | Age | ≥ 70 | 284 (11.1%) | 75.7% | 1.60 | 1.06 | 2.43 |
| | Sex | Female (reference) | 2077 (81.5%) | 67.6% | 1.00 | | |
| | Sex | Male | 471 (18.5%) | 72.8% | 1.18 | 0.80 | 1.73 |
| | Ethnic group | Racially minoritised groups | 72 (2.8%) | 58.3% | 0.56 | 0.28 | 1.09 |
| | Ethnic group | White groups (reference) | 2476 (97.2%) | 68.9% | 1.00 | | |
| | Highest educational qualification | Prefer not to answer/not stated | 102 (4.0%) | 56.9% | 0.27 | 0.10 | 0.74 |
| | Highest educational qualification | Less than University degree or equivalent | 701 (27.5%) | 64.8% | 0.54 | 0.39 | 0.76 |
| | Highest educational qualification | University degree (reference) | 874 (34.3%) | 70.8% | 1.00 | | |
| | Highest educational qualification | Postgraduate degree or higher | 871 (34.2%) | 70.8% | 0.88 | 0.65 | 1.19 |
| | Pre-pandemic employment status | Employed (reference) | 1360 (53.4%) | 66.0% | 1.00 | | |
| | Pre-pandemic employment status | Self-employed | 294 (11.5%) | 68.7% | 0.91 | 0.66 | 1.27 |
| | Pre-pandemic employment status | Unemployed | 11 (0.4%) | 63.6% | 1.03 | 0.26 | 4.07 |
| | Pre-pandemic employment status | Permanently or long-term sick or disabled | 32 (1.3%) | 53.1% | 1.50 | 0.58 | 3.87 |
| | Pre-pandemic employment status | Retired | 568 (22.3%) | 76.4% | 1.35 | 0.91 | 2.01 |
| | Pre-pandemic employment status | Other | 216 (8.5%) | 68.5% | 1.17 | 0.78 | 1.76 |

| | | | | | | | |
|---|---|---|---|---|---|---|---|
| | Pre-pandemic employment status | Unknown | 67 (2.6%) | 62.7% | 0.77 | 0.37 | 1.57 |
| | Local area deprivation | IMD Decile 1-3 (most deprived 30%) | 299 (11.7%) | 62.2% | 0.98 | 0.66 | 1.45 |
| | Local area deprivation | IMD Decile 4-7 (reference) | 999 (39.2%) | 67.1% | 1.00 | | |
| | Local area deprivation | IMD Decile 8-10 (least deprived 30%) | 1250 (49.1%) | 71.4% | 1.25 | 0.96 | 1.62 |
| | Region | East Midlands | 151 (5.9%) | 60.3% | 0.35 | 0.19 | 0.65 |
| | Region | East of England | 275 (10.8%) | 68.4% | 0.73 | 0.43 | 1.23 |
| | Region | London (reference) | 475 (18.6%) | 71.2% | 1.00 | | |
| | Region | North East | 78 (3.1%) | 69.2% | 0.84 | 0.44 | 1.59 |
| | Region | North West | 274 (10.8%) | 65.0% | 0.85 | 0.55 | 1.31 |
| | Region | Scotland & Northern Ireland | 119 (4.7%) | 63.0% | 0.50 | 0.29 | 0.88 |
| | Region | South East | 483 (19.0%) | 72.0% | 1.23 | 0.84 | 1.82 |
| | Region | South West | 253 (9.9%) | 71.1% | 0.94 | 0.60 | 1.47 |
| | Region | Wales | 115 (4.5%) | 63.5% | 1.00 | 0.57 | 1.76 |
| | Region | West Midlands | 158 (6.2%) | 71.5% | 0.71 | 0.41 | 1.25 |
| | Region | Yorkshire and The Humber | 167 (6.6%) | 65.9% | 0.66 | 0.37 | 1.16 |
| **Composite pre-pandemic demographics** | Number of disadvantage indicators (unit: +1) | | | | 0.74 | 0.62 | 0.88 |
| | Number of disadvantage indicators | 0/5 | 299 (11.7%) | 76.3% | 1.56 | 0.96 | 2.53 |
| | Number of disadvantage indicators | 1/5 (reference) | 1436 (56.4%) | 69.6% | 1.00 | | |
| | Number of disadvantage indicators | 2 of 5 | 689 (27.0%) | 65.9% | 0.90 | 0.69 | 1.17 |
| | Number of disadvantage indicators | ≥ 3/5 | 124 (4.9%) | 54.0% | 0.45 | 0.26 | 0.76 |
| | Number of advantage indicators (unit: +1) | | | | 1.26 | 1.08 | 1.47 |
| | Number of advantage indicators | 0/4 | 15 (0.6%) | 53.3% | 0.45 | 0.14 | 1.51 |
| | Number of advantage indicators | 1 of 4 | 739 (29.0%) | 63.6% | 0.63 | 0.47 | 0.84 |
| | Number of advantage indicators | 2/4 (reference) | 1145 (44.9%) | 69.4% | 1.00 | | |
| | Number of advantage indicators | 3 of 4 | 557 (21.9%) | 72.2% | 0.90 | 0.64 | 1.25 |
| | Number of advantage indicators | 4 of 4 | 92 (3.6%) | 79.3% | 1.58 | 0.81 | 3.09 |
| | Number of advantages - number of disadvantages (unit: +1) | | | | 1.18 | 1.07 | 1.30 |
| | Number of advantages - number of disadvantages | ≤ -3 | 12 (0.5%) | 58.3% | 0.67 | 0.19 | 2.31 |

| | | | | | | | |
|---|---|---|---|---|---|---|---|
| | Number of advantages - number of disadvantages | -2 | 115 (4.5%) | 52.2% | 0.38 | 0.21 | 0.67 |
| | Number of advantages - number of disadvantages | -1 | 316 (12.4%) | 62.7% | 0.72 | 0.48 | 1.07 |
| | Number of advantages - number of disadvantages | 0 | 681 (26.7%) | 69.2% | 0.87 | 0.63 | 1.22 |
| | Number of advantages - number of disadvantages | +1 (reference) | 717 (28.1%) | 68.8% | 1.00 | | |
| | Number of advantages - number of disadvantages | 2 | 471 (18.5%) | 71.3% | 1.07 | 0.74 | 1.55 |
| | Number of advantages - number of disadvantages | 3 | 144 (5.7%) | 76.4% | 1.07 | 0.53 | 2.17 |
| | Number of advantages - number of disadvantages | 4 | 92 (3.6%) | 79.3% | 1.69 | 0.85 | 3.36 |
| **Socio-demographics during pandemic** | Pandemic experiences, Lost job/unable to work | No (reference) | 2245 (88.1%) | 71.1% | 1.00 | | |
| | Pandemic experiences, Lost job/unable to work | Yes | 303 (11.9%) | 50.2% | 0.44 | 0.31 | 0.62 |
| | Pandemic experiences, Evicted/lost accommodation | No (reference) | 2531 (99.3%) | 68.6% | 1.00 | | |
| | Pandemic experiences, Evicted/lost accommodation | Yes | 17 (0.7%) | 64.7% | 1.79 | 0.54 | 6.01 |
| | Pandemic experiences, Unable to afford food/bills | No (reference) | 2441 (95.8%) | 69.7% | 1.00 | | |
| | Pandemic experiences, Unable to afford food/bills | Yes | 107 (4.2%) | 43.9% | 0.51 | 0.28 | 0.91 |
| | Pandemic experiences, health & social care access issues (of 5) | None (reference) | 1838 (72.1%) | 72.1% | 1.00 | | |
| | Pandemic experiences, health & social care access issues (of 5) | One | 437 (17.2%) | 63.8% | 0.70 | 0.52 | 0.94 |
| | Pandemic experiences, health & social care access issues (of 5) | Two | 183 (7.2%) | 56.8% | 0.72 | 0.45 | 1.17 |
| | Pandemic experiences, health & social care access issues (of 5) | Three or more | 90 (3.5%) | 43.3% | 0.30 | 0.16 | 0.54 |
| | Pandemic experiences, Lost somebody close due to COVID-19 | No (reference) | 2214 (86.9%) | 70.5% | 1.00 | | |
| | Pandemic experiences, Lost somebody close due to COVID-19 | Yes | 334 (13.1%) | 56.3% | 0.55 | 0.39 | 0.79 |
| | Overall number of negative pandemic experiences (of 10) | None (reference) | 1471 (57.7%) | 75.1% | 1.00 | | |
| | Overall number of negative pandemic experiences (of 10) | One | 624 (24.5%) | 66.3% | 0.68 | 0.52 | 0.90 |

| | | | | | | | |
|---|---|---|---|---|---|---|---|
| | Overall number of negative pandemic experiences (of 10) | Two | 263 (10.3%) | 55.5% | 0.56 | 0.39 | 0.82 |
| | Overall number of negative pandemic experiences (of 10) | Three | 109 (4.3%) | 48.6% | 0.40 | 0.20 | 0.78 |
| | Overall number of negative pandemic experiences (of 10) | Four or more | 81 (3.2%) | 38.3% | 0.26 | 0.14 | 0.48 |
| | Current employment status | Employed (reference) | 1166 (45.8%) | 67.6% | 1.00 | | |
| | Current employment status | Self-employed | 282 (11.1%) | 69.9% | 0.90 | 0.63 | 1.29 |
| | Current employment status | Unemployed | 27 (1.1%) | 29.6% | 0.18 | 0.07 | 0.47 |
| | Current employment status | Permanently or long-term sick or disabled | 62 (2.4%) | 30.6% | 0.25 | 0.11 | 0.56 |
| | Current employment status | Retired | 710 (27.9%) | 74.8% | 1.18 | 0.82 | 1.71 |
| | Current employment status | Other | 231 (9.1%) | 71.4% | 1.09 | 0.72 | 1.66 |
| | Current employment status | Unknown | 70 (2.7%) | 57.1% | 1.01 | 0.32 | 3.18 |
| | Household income | Prefer not to answer/not stated | 387 (15.2%) | 65.4% | 1.14 | 0.71 | 1.83 |
| | Household income | Less than £20,000 | 197 (7.7%) | 51.8% | 0.51 | 0.30 | 0.87 |
| | Household income | £20,000-£29,999 | 229 (9.0%) | 65.5% | 1.07 | 0.67 | 1.71 |
| | Household income | £30,000-£39,999 | 268 (10.5%) | 65.3% | 0.76 | 0.48 | 1.20 |
| | Household income | £40,000-£49,999 (reference) | 303 (11.9%) | 68.0% | 1.00 | | |
| | Household income | £50,000-£74,999 | 475 (18.6%) | 71.2% | 1.09 | 0.69 | 1.72 |
| | Household income | £75,000-£99,999 | 284 (11.1%) | 70.1% | 1.07 | 0.62 | 1.82 |
| | Household income | £100,000 or more | 405 (15.9%) | 80.2% | 2.31 | 1.43 | 3.73 |
| **Pre-pandemic health factors** | Pre-pandemic general health | Poor | 65 (2.6%) | 55.4% | 1.20 | 0.55 | 2.62 |
| | Pre-pandemic general health | Fair | 179 (7.0%) | 54.2% | 0.82 | 0.53 | 1.27 |
| | Pre-pandemic general health | Good | 553 (21.7%) | 63.3% | 0.92 | 0.69 | 1.23 |
| | Pre-pandemic general health | Very good (reference) | 922 (36.2%) | 70.5% | 1.00 | | |
| | Pre-pandemic general health | Excellent | 496 (19.5%) | 76.6% | 1.30 | 0.94 | 1.79 |
| | Pre-pandemic general health | Unknown | 333 (13.1%) | 70.6% | 1.38 | 0.93 | 2.06 |
| | BMI | < 18.5 kg/m^2 | 32 (1.3%) | 68.8% | 0.69 | 0.23 | 2.14 |
| | BMI | 18.5-25 kg/m^2 (reference) | 1112 (43.6%) | 74.6% | 1.00 | | |
| | BMI | 25-30 kg/m^2 | 808 (31.7%) | 66.7% | 0.68 | 0.49 | 0.93 |
| | BMI | ≥ 30 kg/m^2 | 591 (23.2%) | 59.7% | 0.63 | 0.45 | 0.88 |

| | | | | | | | |
|---|---|---|---|---|---|---|---|
| | PRISMA-7 frailty scale | 0-2, below threshold (reference) | 2422 (95.1%) | 69.7% | 1.00 | | |
| | PRISMA-7 frailty scale | 3-7, above threshold | 126 (4.9%) | 48.4% | 0.49 | 0.29 | 0.84 |
| | Physical health conditions | None (reference) | 1958 (76.8%) | 71.0% | 1.00 | | |
| | Physical health conditions | One | 515 (20.2%) | 62.5% | 0.92 | 0.69 | 1.23 |
| | Physical health conditions | Two or more | 75 (2.9%) | 46.7% | 0.44 | 0.22 | 0.89 |
| | Mental health conditions (February 2021) | None (reference) | 1534 (60.2%) | 74.1% | 1.00 | | |
| | Mental health conditions (February 2021) | One | 360 (14.1%) | 60.0% | 0.59 | 0.42 | 0.82 |
| | Mental health conditions (February 2021) | Two | 129 (5.1%) | 59.7% | 0.92 | 0.56 | 1.52 |
| | Mental health conditions (February 2021) | Three or more | 58 (2.3%) | 48.3% | 0.65 | 0.30 | 1.41 |
| | Mental health conditions (February 2021) | Unknown | 467 (18.3%) | 62.3% | 0.71 | 0.52 | 0.98 |
| **COVID-19 illness characteristics** | Infection period | Before 2020-12-08 (pre-vaccination, wild-type dominant) (reference) | 1458 (57.2%) | 64.7% | 1.00 | | |
| | Infection period | 2020-12-08 to 2021-04-25 (alpha-variant dominant) | 268 (10.5%) | 51.9% | 0.63 | 0.44 | 0.90 |
| | Infection period | 2021-04-25 to 2021-12-08 (delta-variant dominant) | 142 (5.6%) | 73.9% | 1.32 | 0.77 | 2.28 |
| | Infection period | After 2021-12-08 (omicron-variant dominant) | 680 (26.7%) | 82.4% | 2.58 | 1.87 | 3.56 |
| | Symptom duration (retrospective self-report) | Less than 2 weeks | 966 (37.9%) | 88.2% | 3.57 | 1.93 | 6.60 |
| | Symptom duration (retrospective self-report) | 2 to 4 weeks | 581 (22.8%) | 74.9% | 1.66 | 0.94 | 2.95 |
| | Symptom duration (retrospective self-report) | 4 to 12 weeks | 330 (13.0%) | 63.0% | 1.15 | 0.63 | 2.08 |
| | Symptom duration (retrospective self-report) | 3 to 6 months (reference) | 150 (5.9%) | 64.0% | 1.00 | | |
| | Symptom duration (retrospective self-report) | 6 to 12 months | 116 (4.6%) | 57.8% | 0.98 | 0.46 | 2.10 |
| | Symptom duration (retrospective self-report) | 12 to 18 months | 76 (3.0%) | 47.4% | 0.81 | 0.32 | 2.08 |
| | Symptom duration (retrospective self-report) | 18 to 24 months | 138 (5.4%) | 19.6% | 0.62 | 0.26 | 1.46 |
| | Symptom duration (retrospective self-report) | 24 months or more | 191 (7.5%) | 14.1% | 0.25 | 0.11 | 0.58 |
| | Symptom duration (prospective logging) | Unknown | 335 (13.1%) | 62.1% | 1.48 | 0.80 | 2.73 |
| | Symptom duration (prospective logging) | Asymptomatic | 188 (7.4%) | 88.8% | 4.64 | 2.02 | 10.67 |
| | Symptom duration (prospective logging) | Less than 2 weeks | 686 (26.9%) | 85.6% | 2.95 | 1.66 | 5.25 |

| | | | | | | | |
|---|---|---|---|---|---|---|---|
| | Symptom duration (prospective logging) | 2 to 4 weeks | 204 (8.0%) | 80.9% | 2.06 | 1.05 | 4.03 |
| | Symptom duration (prospective logging) | 4 to 12 weeks | 576 (22.6%) | 70.1% | 2.11 | 1.25 | 3.58 |
| | Symptom duration (prospective logging) | 3 to 6 months (reference) | 232 (9.1%) | 46.1% | 1.00 | | |
| | Symptom duration (prospective logging) | 6 to 12 months | 146 (5.7%) | 43.2% | 1.04 | 0.53 | 2.07 |
| | Symptom duration (prospective logging) | 12 to 18 months | 132 (5.2%) | 27.3% | 0.64 | 0.29 | 1.43 |
| | Symptom duration (prospective logging) | 18 to 24 months | 32 (1.3%) | 28.1% | 0.48 | 0.13 | 1.71 |
| | Symptom duration (prospective logging) | 24 months or more | 17 (0.7%) | 11.8% | 0.34 | 0.02 | 4.90 |
| | Affected function duration | Able to function as normal | 420 (16.5%) | 81.4% | 0.86 | 0.58 | 1.28 |
| | Affected function duration | Less than 2 weeks (reference) | 915 (35.9%) | 84.4% | 1.00 | | |
| | Affected function duration | 2 to 4 weeks | 294 (11.5%) | 76.2% | 0.90 | 0.56 | 1.44 |
| | Affected function duration | 4 to 12 weeks | 248 (9.7%) | 69.4% | 0.58 | 0.34 | 0.99 |
| | Affected function duration | 3 to 6 months | 170 (6.7%) | 60.0% | 0.48 | 0.29 | 0.81 |
| | Affected function duration | 6 to 12 months | 105 (4.1%) | 58.1% | 0.64 | 0.34 | 1.22 |
| | Affected function duration | 12 to 18 months | 59 (2.3%) | 50.8% | 0.28 | 0.11 | 0.72 |
| | Affected function duration | 18 to 24 months | 147 (5.8%) | 12.9% | 0.11 | 0.05 | 0.24 |
| | Affected function duration | 24 months or more | 181 (7.1%) | 10.5% | 0.08 | 0.04 | 0.18 |
| | New conditions due to COVID-19 | None (reference) | 2279 (89.4%) | 74.2% | 1.00 | | |
| | New conditions due to COVID-19 | One | 110 (4.3%) | 30.9% | 0.65 | 0.32 | 1.33 |
| | New conditions due to COVID-19 | Two | 66 (2.6%) | 22.7% | 0.24 | 0.10 | 0.58 |
| | New conditions due to COVID-19 | Three or more | 68 (2.7%) | 5.9% | 0.07 | 0.02 | 0.24 |
| | Urgent care accessed during COVID-19 illness | No (reference) | 2316 (90.9%) | 71.5% | 1.00 | | |
| | Urgent care accessed during COVID-19 illness | Yes | 232 (9.1%) | 39.7% | 1.32 | 0.78 | 2.24 |
| | Long COVID Diagnosis | No (no self-reported long COVID) | 1446 (56.8%) | 90.0% | 6.13 | 4.11 | 9.15 |
| | Long COVID Diagnosis | No (with self-reported long COVID) (reference) | 762 (29.9%) | 48.8% | 1.00 | | |
| | Long COVID Diagnosis | Yes | 337 (13.2%) | 21.4% | 0.64 | 0.37 | 1.11 |
| **Factors unique to cohort** | First language | English (reference) | 2430 (95.4%) | 68.7% | 1.00 | | |
| | First language | Other | 47 (1.8%) | 78.7% | 3.22 | 1.19 | 8.71 |
| | First language | Prefer not to answer/not stated | 71 (2.8%) | 57.7% | 0.70 | 0.35 | 1.42 |

#### *TwinsUK*

Table S 2. **Extended sample characteristics and results of multivariable logistic regression models, TwinsUK cohort.** Odds ratios and confidence intervals (CI) are presented for multivariable logistic regression models testing association between recovery from COVID-19 and exposure of interest, after adjustment as appropriate from the hypothesised DAG, and weighting for inverse probability of participation.

| | TwinsUK | | | | | | |
|---|---|---|---|---|---|---|---|
| **Domain** | **Variable** | **Category** | **Group size, N (%)** | **COVID-19 recovery (%)** | **Odds ratio** | **95% CI (lower)** | **95% CI (upper)** |
| **Individual pre-pandemic demographics** | Age | 18-39 | 266 (19.9%) | 88.3% | 1.69 | 0.99 | 2.88 |
| | Age | 40-49 | 195 (14.6%) | 79.0% | 0.92 | 0.54 | 1.56 |
| | Age | 50-59 (reference) | 338 (25.3%) | 78.1% | 1.00 | | |
| | Age | 60-69 | 294 (22.0%) | 79.3% | 1.17 | 0.75 | 1.81 |
| | Age | ≥ 70 | 241 (18.1%) | 79.7% | 1.03 | 0.66 | 1.62 |
| | Sex | Female (reference) | 1153 (86.4%) | 79.7% | 1.00 | | |
| | Sex | Male | 181 (13.6%) | 87.8% | 1.91 | 1.12 | 3.25 |
| | Ethnic group | Racially minoritised groups | 52 (3.9%) | 73.1% | 0.54 | 0.24 | 1.23 |
| | Ethnic group | White groups (reference) | 1282 (96.1%) | 81.1% | 1.00 | | |
| | Highest educational qualification | Unknown | 53 (4.0%) | 66.0% | 0.36 | 0.18 | 0.71 |
| | Highest educational qualification | Less than University degree or equivalent | 680 (51.0%) | 78.4% | 0.69 | 0.48 | 1.01 |
| | Highest educational qualification | University degree (reference) | 386 (28.9%) | 84.5% | 1.00 | | |
| | Highest educational qualification | Postgraduate degree or higher | 215 (16.1%) | 85.6% | 1.04 | 0.63 | 1.70 |
| | Pre-pandemic employment status | Employed (reference) | 689 (51.6%) | 82.3% | 1.00 | | |
| | Pre-pandemic employment status | Self-employed | 119 (8.9%) | 80.7% | 0.99 | 0.52 | 1.89 |
| | Pre-pandemic employment status | Unemployed | 9 (0.7%) | 100.0% | | | |
| | Pre-pandemic employment status | Permanently or long-term sick or disabled | 18 (1.3%) | 44.4% | 0.31 | 0.09 | 1.08 |
| | Pre-pandemic employment status | Retired | 279 (20.9%) | 80.3% | 1.05 | 0.57 | 1.91 |
| | Pre-pandemic employment status | Other | 124 (9.3%) | 77.4% | 0.77 | 0.41 | 1.44 |
| | Pre-pandemic employment status | Unknown | 96 (7.2%) | 81.3% | 1.76 | 0.81 | 3.79 |

| | | | | | | | |
|---|---|---|---|---|---|---|---|
| | Local area deprivation | IMD Decile 1-3 (most deprived 30%) | 178 (13.3%) | 78.7% | 1.07 | 0.62 | 1.84 |
| | Local area deprivation | IMD Decile 4-7 (reference) | 533 (40.0%) | 81.1% | 1.00 | | |
| | Local area deprivation | IMD Decile 8-10 (least deprived 30%) | 623 (46.7%) | 81.2% | 1.28 | 0.89 | 1.85 |
| | Region | East Midlands | 69 (5.2%) | 72.5% | 0.46 | 0.22 | 0.98 |
| | Region | East of England | 174 (13.0%) | 83.9% | 1.16 | 0.59 | 2.26 |
| | Region | London (reference) | 266 (19.9%) | 83.1% | 1.00 | | |
| | Region | North East | 32 (2.4%) | 78.1% | 0.51 | 0.16 | 1.56 |
| | Region | North West | 93 (7.0%) | 86.0% | 1.93 | 0.77 | 4.86 |
| | Region | Scotland & Northern Ireland | 45 (3.4%) | 73.3% | 0.45 | 0.17 | 1.16 |
| | Region | South East | 310 (23.2%) | 80.0% | 0.86 | 0.51 | 1.46 |
| | Region | South West | 142 (10.6%) | 81.0% | 1.07 | 0.55 | 2.07 |
| | Region | Wales | 47 (3.5%) | 70.2% | 0.55 | 0.24 | 1.26 |
| | Region | West Midlands | 79 (5.9%) | 82.3% | 0.88 | 0.40 | 1.95 |
| | Region | Yorkshire and The Humber | 77 (5.8%) | 80.5% | 1.74 | 0.77 | 3.93 |
| **Composite pre-pandemic demographics** | Number of disadvantage indicators (unit: +1) | | | | 0.79 | 0.64 | 0.98 |
| | Number of disadvantage indicators | 0/5 | 81 (6.1%) | 90.1% | 2.95 | 1.20 | 7.24 |
| | Number of disadvantage indicators | 1/5 (reference) | 514 (38.5%) | 82.7% | 1.00 | | |
| | Number of disadvantage indicators | 2 of 5 | 620 (46.5%) | 79.7% | 0.93 | 0.63 | 1.37 |
| | Number of disadvantage indicators | ≥ 3/5 | 119 (8.9%) | 72.3% | 0.80 | 0.46 | 1.40 |
| | Number of advantage indicators (unit: +1) | | | | 1.36 | 1.09 | 1.70 |
| | Number of advantage indicators | 0/4 | 39 (2.9%) | 82.1% | 1.03 | 0.35 | 3.02 |
| | Number of advantage indicators | 1/4 (reference) | 518 (38.8%) | 78.8% | 1.00 | | |
| | Number of advantage indicators | 2 of 4 | 592 (44.4%) | 79.9% | 1.21 | 0.84 | 1.73 |
| | Number of advantage indicators | ≥ 3/4 | 185 (13.9%) | 89.2% | 2.78 | 1.55 | 5.01 |
| | Number of advantages - Number of disadvantages (unit: +1) | | | | 1.18 | 1.05 | 1.34 |
| | Number of advantages - Number of disadvantages | ≤ -3 | 30 (2.2%) | 76.7% | 1.33 | 0.52 | 3.42 |
| | Number of advantages - Number of disadvantages | -2 | 98 (7.3%) | 73.5% | 0.66 | 0.31 | 1.41 |

| | | | | | | | |
|---|---|---|---|---|---|---|---|
| | Number of advantages - Number of disadvantages | -1 | 298 (22.3%) | 78.9% | 0.93 | 0.55 | 1.56 |
| | Number of advantages - Number of disadvantages | 0 | 448 (33.6%) | 80.4% | 0.97 | 0.58 | 1.61 |
| | Number of advantages - Number of disadvantages | +1 (reference) | 257 (19.3%) | 80.5% | 1.00 | | |
| | Number of advantages - Number of disadvantages | 2 | 141 (10.6%) | 88.7% | 2.00 | 1.01 | 3.99 |
| | Number of advantages - Number of disadvantages | ≥ +3 | 62 (4.6%) | 90.3% | 3.43 | 1.18 | 9.95 |
| **Socio-demographics during pandemic** | Pandemic experiences, Lost job/unable to work | No (reference) | 1120 (84.0%) | 81.7% | 1.00 | | |
| | Pandemic experiences, Lost job/unable to work | Yes | 206 (15.4%) | 76.7% | 0.52 | 0.31 | 0.87 |
| | Pandemic experiences, Evicted/lost accommodation | No (reference) | 1313 (98.4%) | 80.7% | 1.00 | | |
| | Pandemic experiences, Evicted/lost accommodation | Yes | 14 (1.0%) | 92.9% | 2.66 | 0.31 | 22.77 |
| | Pandemic experiences, Unable to afford food/bills | No (reference) | 1206 (90.4%) | 82.2% | 1.00 | | |
| | Pandemic experiences, Unable to afford food/bills | Yes | 121 (9.1%) | 67.8% | 0.40 | 0.24 | 0.65 |
| | Pandemic experiences, health & social care access issues (of 5) | None (reference) | 759 (56.9%) | 83.8% | 1.00 | | |
| | Pandemic experiences, health & social care access issues (of 5) | One | 360 (27.0%) | 80.8% | 0.98 | 0.65 | 1.45 |
| | Pandemic experiences, health & social care access issues (of 5) | Two | 141 (10.6%) | 71.6% | 0.62 | 0.37 | 1.03 |
| | Pandemic experiences, health & social care access issues (of 5) | Three or more | 69 (5.2%) | 68.1% | 0.40 | 0.21 | 0.76 |
| | Pandemic experiences, Lost somebody close due to COVID-19/Change in relationship status | No (reference) | 1110 (83.2%) | 81.8% | 1.00 | | |
| | Pandemic experiences, Lost somebody close due to COVID-19/Change in relationship status | Yes | 217 (16.3%) | 76.0% | 0.66 | 0.43 | 1.01 |
| | Overall number of negative pandemic experiences (of 11) | None (reference) | 574 (43.0%) | 84.7% | 1.00 | | |
| | Overall number of negative pandemic experiences (of 11) | One | 435 (32.6%) | 81.6% | 0.79 | 0.52 | 1.20 |
| | Overall number of negative pandemic experiences (of 11) | Two | 172 (12.9%) | 79.7% | 0.96 | 0.57 | 1.62 |
| | Overall number of negative pandemic experiences (of 11) | Three | 84 (6.3%) | 69.0% | 0.43 | 0.23 | 0.79 |

| | | | | | | | |
|---|---|---|---|---|---|---|---|
| | Overall number of negative pandemic experiences (of 11) | Four or more | 64 (4.8%) | 60.9% | 0.21 | 0.11 | 0.41 |
| | Current employment status | Employed (reference) | 698 (52.3%) | 83.2% | 1.00 | | |
| | Current employment status | Self-employed | 109 (8.2%) | 81.7% | 1.05 | 0.53 | 2.07 |
| | Current employment status | Unemployed | 11 (0.8%) | 72.7% | 0.53 | 0.10 | 2.76 |
| | Current employment status | Permanently or long-term sick or disabled | 22 (1.6%) | 31.8% | 0.14 | 0.04 | 0.43 |
| | Current employment status | Retired | 343 (25.7%) | 80.2% | 0.95 | 0.53 | 1.73 |
| | Current employment status | Other | 105 (7.9%) | 76.2% | 0.76 | 0.40 | 1.44 |
| | Current employment status | Unknown | 46 (3.4%) | 82.6% | 2.06 | 0.71 | 5.96 |
| | Household income (latest) | Prefer not to answer | 151 (11.3%) | 76.2% | 0.74 | 0.38 | 1.42 |
| | Household income (latest) | Less than £20,000 | 133 (10.0%) | 75.9% | 1.08 | 0.54 | 2.17 |
| | Household income (latest) | £20,000-£29,999 | 162 (12.1%) | 84.0% | 1.36 | 0.69 | 2.69 |
| | Household income (latest) | £30,000-£39,999 | 135 (10.1%) | 82.2% | 1.25 | 0.62 | 2.55 |
| | Household income (latest) | £40,000-£49,999 (reference) | 147 (11.0%) | 81.0% | 1.00 | | |
| | Household income (latest) | £50,000-£74,999 | 227 (17.0%) | 80.6% | 0.86 | 0.47 | 1.57 |
| | Household income (latest) | £75,000-£99,999 | 121 (9.1%) | 87.6% | 1.28 | 0.58 | 2.79 |
| | Household income (latest) | £100,000 or more | 155 (11.6%) | 86.5% | 1.17 | 0.58 | 2.39 |
| | Household income (latest) | Unknown | 103 (7.7%) | 70.9% | 0.59 | 0.26 | 1.32 |
| **Pre-pandemic health factors** | Pre-pandemic general health | Poor | 17 (1.3%) | 35.3% | 0.21 | 0.06 | 0.71 |
| | Pre-pandemic general health | Fair | 80 (6.0%) | 68.8% | 0.78 | 0.36 | 1.67 |
| | Pre-pandemic general health | Good | 295 (22.1%) | 77.6% | 0.80 | 0.52 | 1.23 |
| | Pre-pandemic general health | Very good (reference) | 458 (34.3%) | 82.5% | 1.00 | | |
| | Pre-pandemic general health | Excellent | 312 (23.4%) | 88.1% | 1.75 | 1.02 | 3.02 |
| | Pre-pandemic general health | Unknown | 172 (12.9%) | 78.5% | 0.81 | 0.38 | 1.69 |
| | BMI | < 18.5 kg/m^2 | 35 (2.6%) | 94.3% | 3.42 | 0.74 | 15.84 |
| | BMI | 18.5-25 kg/m^2 (reference) | 580 (43.5%) | 83.1% | 1.00 | | |
| | BMI | 25-30 kg/m^2 | 359 (26.9%) | 80.2% | 1.13 | 0.75 | 1.71 |
| | BMI | ≥ 30 kg/m^2 | 192 (14.4%) | 76.0% | 1.28 | 0.73 | 2.25 |
| | BMI | Unknown | 168 (12.6%) | 76.8% | 0.47 | 0.18 | 1.20 |

| | | | | | | | |
|---|---|---|---|---|---|---|---|
| | PRISMA-7 frailty scale | 0-2, below threshold (reference) | 802 (60.1%) | 82.0% | 1.00 | | |
| | PRISMA-7 frailty scale | 3-7, above threshold | 43 (3.2%) | 67.4% | 0.62 | 0.24 | 1.57 |
| | PRISMA-7 frailty scale | Unknown | 489 (36.7%) | 80.0% | 1.11 | 0.71 | 1.71 |
| | Physical health conditions | None (reference) | 703 (52.7%) | 82.2% | 1.00 | | |
| | Physical health conditions | One | 304 (22.8%) | 84.2% | 1.43 | 0.93 | 2.20 |
| | Physical health conditions | Two or more | 90 (6.7%) | 66.7% | 0.64 | 0.35 | 1.15 |
| | Physical health conditions | Unknown | 237 (17.8%) | 77.6% | 2.08 | 0.88 | 4.92 |
| | Mental health conditions (pre-pandemic) | None (reference) | 804 (60.3%) | 83.8% | 1.00 | | |
| | Mental health conditions (pre-pandemic) | One | 185 (13.9%) | 77.3% | 0.70 | 0.45 | 1.10 |
| | Mental health conditions (pre-pandemic) | Two or more | 108 (8.1%) | 71.3% | 0.49 | 0.28 | 0.85 |
| **COVID-19 illness characteristics** | Infection period | Before 2020-12-08 (pre-vaccination, wild-type dominant) (reference) | 677 (50.7%) | 78.0% | 1.00 | | |
| | Infection period | 2020-12-08 to 2021-04-25 (alpha-variant dominant) | 148 (11.1%) | 77.7% | 0.96 | 0.53 | 1.75 |
| | Infection period | 2021-04-25 to 2021-12-08 (delta-variant dominant) | 205 (15.4%) | 77.1% | 0.85 | 0.53 | 1.37 |
| | Infection period | After 2021-12-08 (omicron-variant dominant) | 304 (22.8%) | 91.1% | 2.87 | 1.77 | 4.65 |
| | Symptom duration (retrospective self-report) | Less than 2 weeks (including asymptomatic) | 603 (45.2%) | 93.4% | 2.70 | 1.56 | 4.69 |
| | Symptom duration (retrospective self-report) | 2 to 4 weeks (reference) | 289 (21.7%) | 85.5% | 1.00 | | |
| | Symptom duration (retrospective self-report) | 4 to 12 weeks | 217 (16.3%) | 76.5% | 0.52 | 0.28 | 0.97 |
| | Symptom duration (retrospective self-report) | 12 or more weeks | 92 (6.9%) | 56.5% | 0.17 | 0.08 | 0.34 |
| | Symptom duration (retrospective self-report) | 3 to 12 months | 69 (5.2%) | 53.6% | 0.20 | 0.10 | 0.42 |
| | Symptom duration (retrospective self-report) | 12 or more months | 64 (4.8%) | 20.3% | 0.03 | 0.01 | 0.06 |
| | Affected function duration | Unknown | 292 (21.9%) | 88.0% | 0.59 | 0.26 | 1.33 |
| | Affected function duration | N/A - No symptoms | 77 (5.8%) | 85.7% | 0.64 | 0.26 | 1.57 |
| | Affected function duration | Able to function as normal | 224 (16.8%) | 86.6% | 1.10 | 0.59 | 2.05 |
| | Affected function duration | Less than 2 weeks (reference) | 430 (32.2%) | 85.6% | 1.00 | | |
| | Affected function duration | 2 to 4 weeks | 122 (9.1%) | 72.1% | 1.01 | 0.53 | 1.93 |
| | Affected function duration | 4 to 12 weeks | 80 (6.0%) | 67.5% | 0.69 | 0.32 | 1.46 |
| | Affected function duration | 12 or more weeks | 79 (5.9%) | 34.2% | 0.50 | 0.23 | 1.11 |

| | | | | | | | |
|---|---|---|---|---|---|---|---|
| | Hospitalised during COVID-19 illness | No (reference) | 1293 (96.9%) | 81.9% | 1.00 | | |
| | Hospitalised during COVID-19 illness | Yes | 37 (2.8%) | 51.4% | 1.37 | 0.47 | 4.02 |
| | Long COVID Diagnosis | No (self-reported long COVID status unknown) | 76 (5.7%) | 68.4% | 1.82 | 0.79 | 4.21 |
| | Long COVID Diagnosis | No (no self-reported long COVID) | 796 (59.7%) | 92.8% | 10.05 | 5.94 | 17.01 |
| | Long COVID Diagnosis | No (with self-reported long COVID) (reference) | 184 (13.8%) | 45.7% | 1.00 | | |
| | Long COVID Diagnosis | Yes | 47 (3.5%) | 23.4% | 0.56 | 0.20 | 1.63 |
| **Factors unique to cohort** | Housing tenure | Owned outright | 387 (29.0%) | 79.6% | 1.00 | | |
| | Housing tenure | Owned with mortgage | 302 (22.6%) | 80.8% | 1.05 | 0.64 | 1.73 |
| | Housing tenure | Rented | 102 (7.6%) | 84.3% | 1.65 | 0.77 | 3.51 |
| | Housing tenure | Other | 55 (4.1%) | 90.9% | 2.54 | 0.81 | 7.98 |
| | Housing, trouble with vermin, damp or mould | No (reference) | 748 (56.1%) | 82.9% | 1.00 | | |
| | Housing, trouble with vermin, damp or mould | Yes | 98 (7.3%) | 69.4% | 0.53 | 0.31 | 0.91 |
| | Household income (pre-pandemic) | Prefer not to answer | 143 (10.7%) | 73.4% | 0.62 | 0.31 | 1.25 |
| | Household income (pre-pandemic) | Less than £20,000 | 102 (7.6%) | 82.4% | 1.94 | 0.87 | 4.36 |
| | Household income (pre-pandemic) | £20,000-£29,999 | 114 (8.5%) | 82.5% | 1.18 | 0.54 | 2.56 |
| | Household income (pre-pandemic) | £30,000-£39,999 | 106 (7.9%) | 82.1% | 1.23 | 0.58 | 2.63 |
| | Household income (pre-pandemic) | £40,000-£49,999 (reference) | 115 (8.6%) | 82.6% | 1.00 | | |
| | Household income (pre-pandemic) | £50,000-£74,999 | 185 (13.9%) | 85.4% | 1.12 | 0.55 | 2.26 |
| | Household income (pre-pandemic) | £75,000-£99,999 | 98 (7.3%) | 85.7% | 0.87 | 0.37 | 2.06 |
| | Household income (pre-pandemic) | £100,000 or more | 112 (8.4%) | 83.0% | 0.79 | 0.36 | 1.74 |
| | Household income (pre-pandemic) | Unknown | 359 (26.9%) | 77.4% | 0.58 | 0.29 | 1.16 |
| | Credit/benefit claims before pandemic | None (reference) | 1072 (80.4%) | 82.6% | 1.00 | | |
| | Credit/benefit claims before pandemic | One or more | 91 (6.8%) | 68.1% | 0.57 | 0.31 | 1.04 |
| | Credit/benefit claims before pandemic | Unknown | 171 (12.8%) | 76.0% | 0.62 | 0.35 | 1.10 |
| | Credit/benefit claims during pandemic | None (reference) | 1080 (81.0%) | 82.6% | 1.00 | | |
| | Credit/benefit claims during pandemic | One or more | 209 (15.7%) | 74.2% | 0.60 | 0.38 | 0.95 |
| | Credit/benefit claims during pandemic | Unknown | 45 (3.4%) | 68.9% | 0.36 | 0.15 | 0.87 |

| | | | | | | | |
|---|---|---|---|---|---|---|---|
| | Count (of 16) significant stressors (April-August 2020) | None (reference) | 530 (39.7%) | 84.0% | 1.00 | | |
| | Count (of 16) significant stressors (April-August 2020) | One | 151 (11.3%) | 80.1% | 0.56 | 0.33 | 0.97 |
| | Count (of 16) significant stressors (April-August 2020) | Two | 165 (12.4%) | 80.0% | 0.75 | 0.44 | 1.27 |
| | Count (of 16) significant stressors (April-August 2020) | Three or more | 308 (23.1%) | 76.6% | 0.69 | 0.44 | 1.07 |
| | Count (of 16) significant stressors (April-August 2020) | Unknown | 180 (13.5%) | 80.0% | 0.65 | 0.34 | 1.23 |
| | Number of caring responsibilities (of 8) | None (reference) | 449 (33.7%) | 82.4% | 1.00 | | |
| | Number of caring responsibilities (of 8) | One | 335 (25.1%) | 81.5% | 1.10 | 0.69 | 1.77 |
| | Number of caring responsibilities (of 8) | Two | 258 (19.3%) | 79.5% | 0.99 | 0.60 | 1.61 |
| | Number of caring responsibilities (of 8) | Three | 69 (5.2%) | 79.7% | 0.78 | 0.39 | 1.59 |
| | Number of caring responsibilities (of 8) | Four or more | 84 (6.3%) | 77.4% | 0.85 | 0.43 | 1.68 |

#### Mediation model logistic regression results

Table S 3. **Results of models estimating association between COVID-19 recovery and socio-demographic composite variables, including potential mediators.** Models adjusted for age group, region and pre-pandemic health factors, and weighted for inverse probability of participation in questionnaires.

| | | COVID Symptom Study Biobank (CSSB) | | | | TwinsUK | | | |
|---|---|---|---|---|---|---|---|---|---|
| **Socio-demographic composite variable** | **Mediator domain** | **Mediators** | **Odds ratio*** | **95% CI (lower)** | **95% CI (upper)** | **Mediators** | **Odds ratio** | **95% CI (lower)** | **95% CI (upper)** |
| Number of disadvantage indicators (unit: +1) | | None | 0.74 | 0.62 | 0.88 | None | 0.79 | 0.64 | 0.98 |
| | COVID-19 illness | COVID-19 infection period, Urgent care, New conditions | 0.76 | 0.62 | 0.93 | COVID-19 infection period, Hospitalised | 0.79 | 0.64 | 0.98 |
| | | COVID-19 infection period, Urgent care, New conditions, Symptom duration (retrospective) | 0.85 | 0.70 | 1.04 | COVID-19 infection period, Hospitalised, Symptom duration (retrospective) | 0.81 | 0.63 | 1.03 |
| | | COVID-19 infection period, Urgent care, New conditions, Symptom duration (prospective) | 0.76 | 0.62 | 0.93 | COVID-19 infection period, Hospitalised, Long COVID status | 0.79 | 0.62 | 1.01 |
| | | COVID-19 infection period, Urgent care, New conditions, Long COVID status | 0.81 | 0.65 | 1.00 | | | | |
| | Adverse pandemic experiences | Lost job/Unable to work | 0.75 | 0.62 | 0.90 | Lost job/Unable to work | 0.82 | 0.66 | 1.01 |
| | | Unable to afford food/bills | 0.75 | 0.63 | 0.90 | Unable to afford food/bills | 0.83 | 0.67 | 1.02 |
| | | Difficulties accessing health & social care | 0.75 | 0.63 | 0.90 | Difficulties accessing health & social care | 0.81 | 0.65 | 1.00 |
| | | Loss of somebody close due to COVID-19 | 0.75 | 0.63 | 0.90 | Loss of somebody close due to COVID-19 | 0.81 | 0.65 | 1.01 |
| | | Count of adverse pandemic experiences | 0.76 | 0.63 | 0.92 | Count of adverse pandemic experiences | 0.82 | 0.66 | 1.01 |
| Number of advantage indicators (unit: +1) | | None | 1.26 | 1.08 | 1.47 | None | 1.36 | 1.09 | 1.70 |
| | COVID-19 illness | COVID-19 infection period, Urgent care, New conditions | 1.25 | 1.05 | 1.48 | COVID-19 infection period, Hospitalised | 1.34 | 1.06 | 1.68 |
| | | COVID-19 infection period, Urgent care, New conditions, Symptom duration (retrospective) | 1.14 | 0.96 | 1.37 | COVID-19 infection period, Hospitalised, Symptom duration (retrospective) | 1.38 | 1.06 | 1.80 |
| | | COVID-19 infection period, Urgent care, New conditions, Symptom duration (prospective) | 1.25 | 1.05 | 1.49 | COVID-19 infection period, Hospitalised, Long COVID status | 1.33 | 1.02 | 1.73 |
| | | COVID-19 infection period, Urgent care, New conditions, Long COVID status | 1.22 | 1.01 | 1.48 | | | | |
| | | Lost job/Unable to work | 1.26 | 1.08 | 1.47 | Lost job/Unable to work | 1.32 | 1.05 | 1.65 |

| | | | | | | | | | |
|---|---|---|---|---|---|---|---|---|---|
| | Adverse pandemic experiences | Unable to afford food/bills | 1.25 | 1.07 | 1.45 | Unable to afford food/bills | 1.31 | 1.05 | 1.64 |
| | | Difficulties accessing health & social care | 1.22 | 1.05 | 1.43 | Difficulties accessing health & social care | 1.33 | 1.06 | 1.66 |
| | | Loss of somebody close due to COVID-19 | 1.24 | 1.07 | 1.45 | Loss of somebody close due to COVID-19 | 1.33 | 1.06 | 1.67 |
| | | Count of adverse pandemic experiences | 1.21 | 1.04 | 1.41 | Count of adverse pandemic experiences | 1.28 | 1.02 | 1.61 |
| Number of advantages – number of disadvantages (unit: +1) | | None | 1.18 | 1.07 | 1.30 | None | 1.18 | 1.05 | 1.34 |
| | COVID-19 illness | COVID-19 infection period, Urgent care, New conditions | 1.17 | 1.05 | 1.30 | COVID-19 infection period, Hospitalised | 1.18 | 1.04 | 1.33 |
| | | COVID-19 infection period, Urgent care, New conditions, Symptom duration (retrospective) | 1.10 | 0.98 | 1.22 | COVID-19 infection period, Hospitalised, Symptom duration (retrospective) | 1.18 | 1.02 | 1.36 |
| | | COVID-19 infection period, Urgent care, New conditions, Symptom duration (prospective) | 1.17 | 1.05 | 1.30 | COVID-19 infection period, Hospitalised, Long COVID status | 1.17 | 1.02 | 1.35 |
| | | COVID-19 infection period, Urgent care, New conditions, Long COVID status | 1.14 | 1.01 | 1.28 | | | | |
| | Adverse pandemic experiences | Lost job/Unable to work | 1.18 | 1.07 | 1.30 | Lost job/Unable to work | 1.16 | 1.03 | 1.32 |
| | | Unable to afford food/bills | 1.17 | 1.06 | 1.29 | Unable to afford food/bills | 1.16 | 1.02 | 1.31 |
| | | Difficulties accessing health & social care | 1.17 | 1.06 | 1.28 | Difficulties accessing health & social care | 1.17 | 1.03 | 1.32 |
| | | Loss of somebody close due to COVID-19 | 1.17 | 1.06 | 1.29 | Loss of somebody close due to COVID-19 | 1.17 | 1.03 | 1.32 |
| | | Count of adverse pandemic experiences | 1.16 | 1.05 | 1.28 | Count of adverse pandemic experiences | 1.15 | 1.02 | 1.30 |

#### Work and social adjustment scale (WSAS) by COVID-19 recovery and long COVID diagnosis status

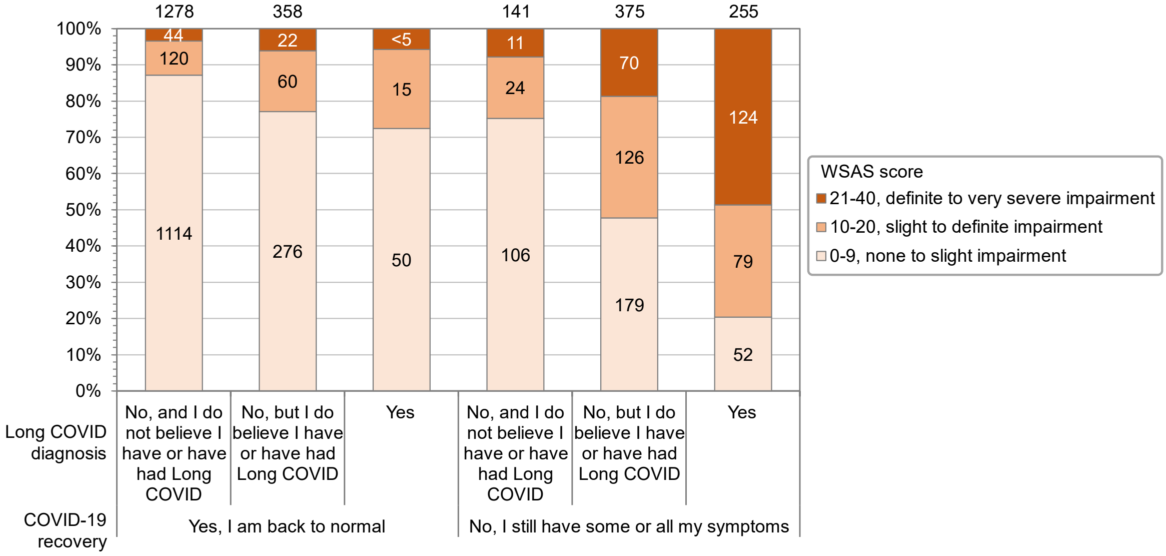

Figure S 3. **Work and social adjustment scale (WSAS) by COVID-19 recovery and long COVID diagnosis status among CSSB participants.** Results shown for all CSSB participants with self-reported COVID-19 infection who completed the WSAS assessment. Data labels show the sample sizes, including totals above each bar. Sample sizes < 5 and associated totals are suppressed.

#### Associations with health characteristics

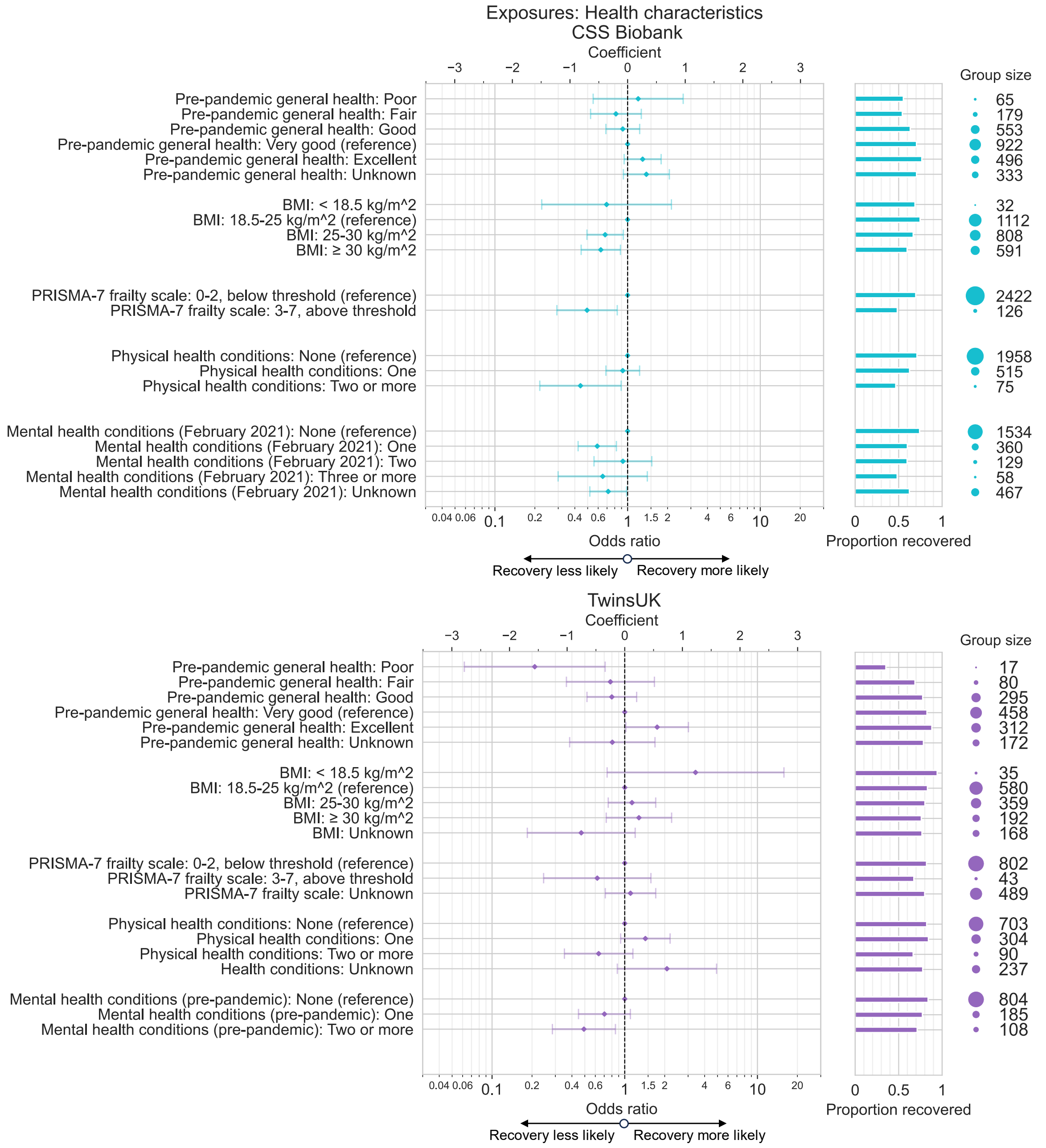

Figure S 4. **Associations between health characteristics and recovery from COVID-19 in CSS Biobank and TwinsUK cohorts**. Odds ratio and 95% confidence intervals from logistic regression models testing association between recovery from COVID-19 and various health-related exposure variables, among individuals with self-reported COVID-19 infection. Results for each exposure variable originate from distinct models, including age, sex, ethnic group, education, pre-pandemic socio-demographics, and other concurrent health characteristics as potential confounding factors.

#### Association between recovery and number of pre-pandemic social advantages or disadvantages as categorical variable

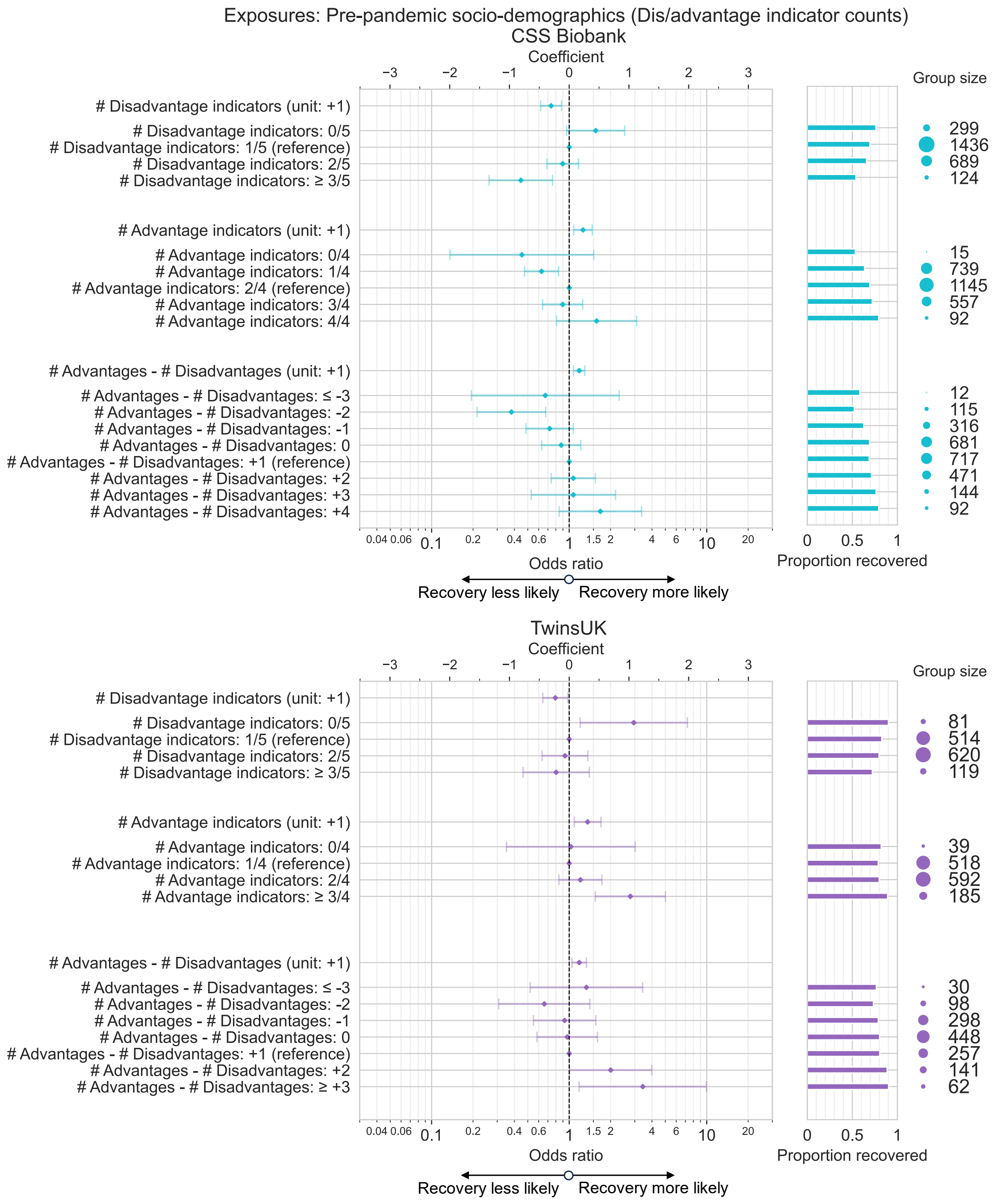

Figure S 5. **Associations between pre-pandemic indicators of socio-demographic advantage and/or disadvantage and recovery from COVID-19 in CSS Biobank and TwinsUK cohorts**. Odds ratio and 95% confidence intervals from logistic regression models testing association between recovery from COVID-19 and composite measures of pre-pandemic of social advantage and disadvantage, among individuals with self-reported COVID-19 infection. Composite indicators were generated from sex, ethnic group, highest educational qualification, local area deprivation and pre-pandemic employment status. Models for both cohorts adjusted for age group, region, pre-pandemic self-rated general health, BMI, frailty and number of physical health conditions. TwinsUK models additionally adjusted for number of mental health conditions.

### Associations with COVID-19 illness characteristics

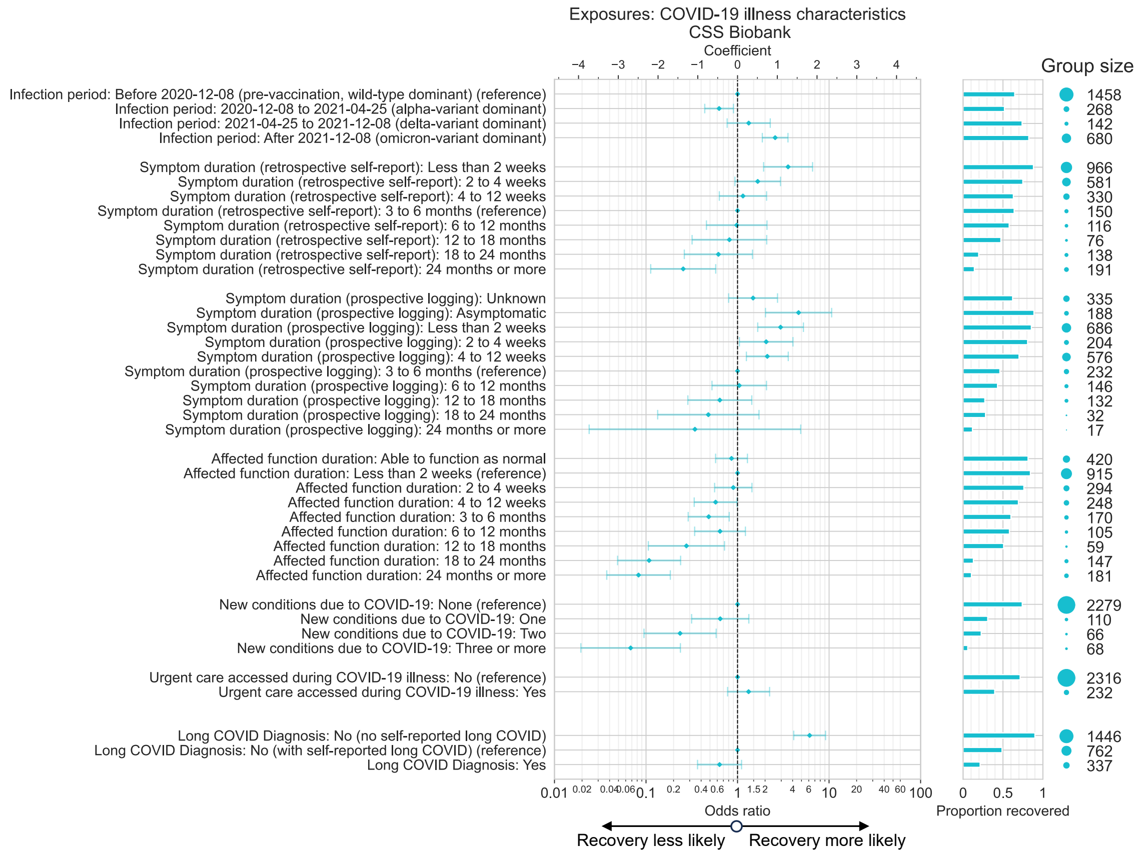

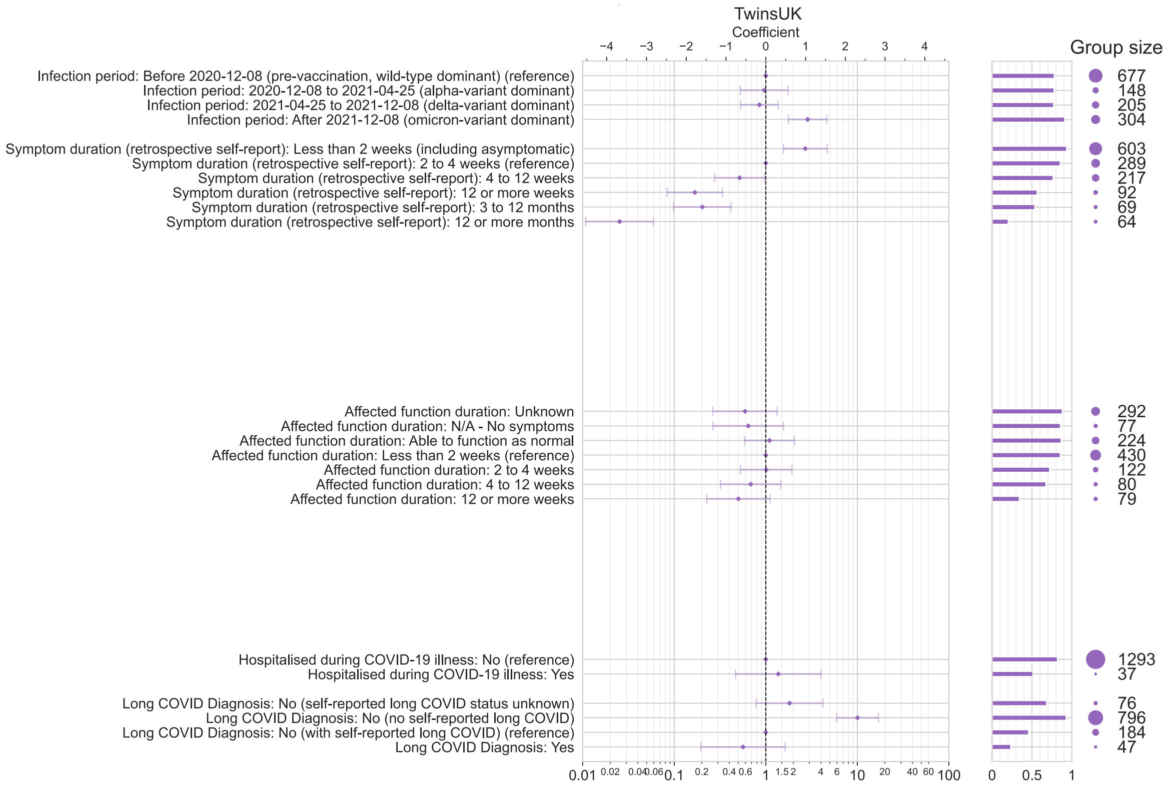

Figure S 6. **Associations between COVID-19 illness characteristics and recovery from COVID-19 in CSS Biobank and TwinsUK cohorts**. Odds ratio and 95% confidence intervals from logistic regression models testing association between recovery from COVID-19 and various COVID-19 illness-related exposure variables, among individuals with self-reported COVID-19 infection. Results for each exposure variable originate from distinct models, including age, sex, ethnic group, education, pre-pandemic socio-demographics, health characteristics and other COVID-19 illness factors as potential confounding factors.

#### Sensitivity analysis stratified by sex

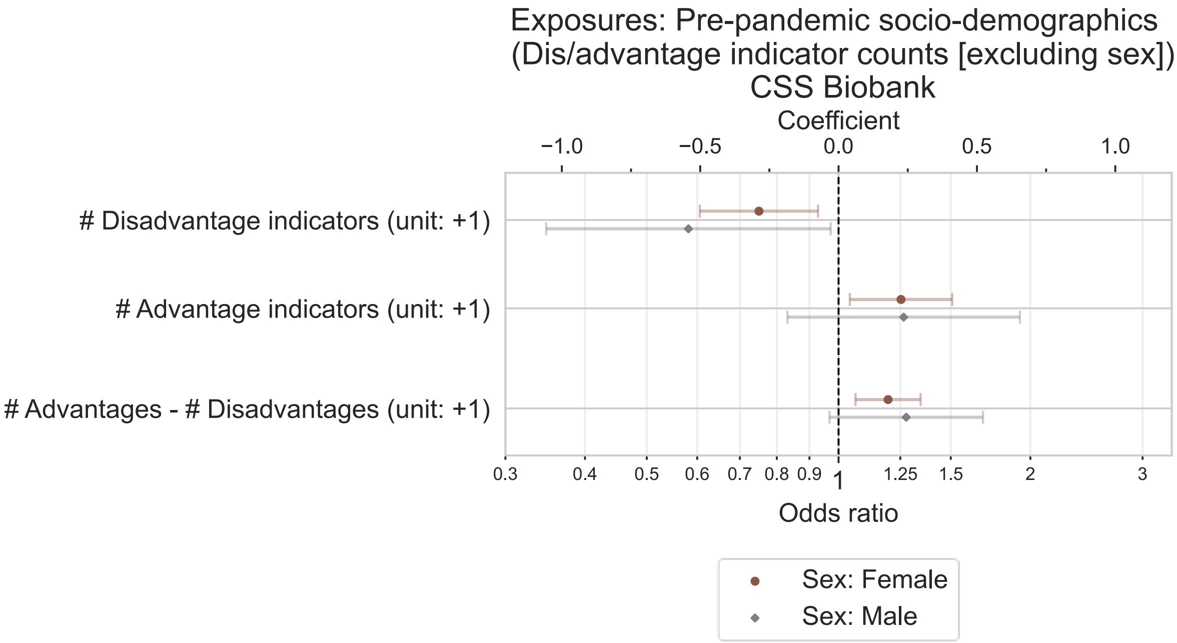

Figure S 7. **Sex-stratified models testing associations between pre-pandemic socio-demographic advantage and/or disadvantage and recovery from COVID-19 in CSSB cohort**. Odds ratio and 95% confidence intervals from logistic regression models testing association between recovery from COVID-19 and composite measures of pre-pandemic of social advantage and disadvantage, among individuals with self-reported COVID-19 infection. Composite indicators were generated from ethnic group, highest educational qualification, local area deprivation and pre-pandemic employment status. Models adjusted for age group, region, pre-pandemic self-rated general health, BMI, frailty and number of physical health conditions.

#### Associations with variables unique to cohort

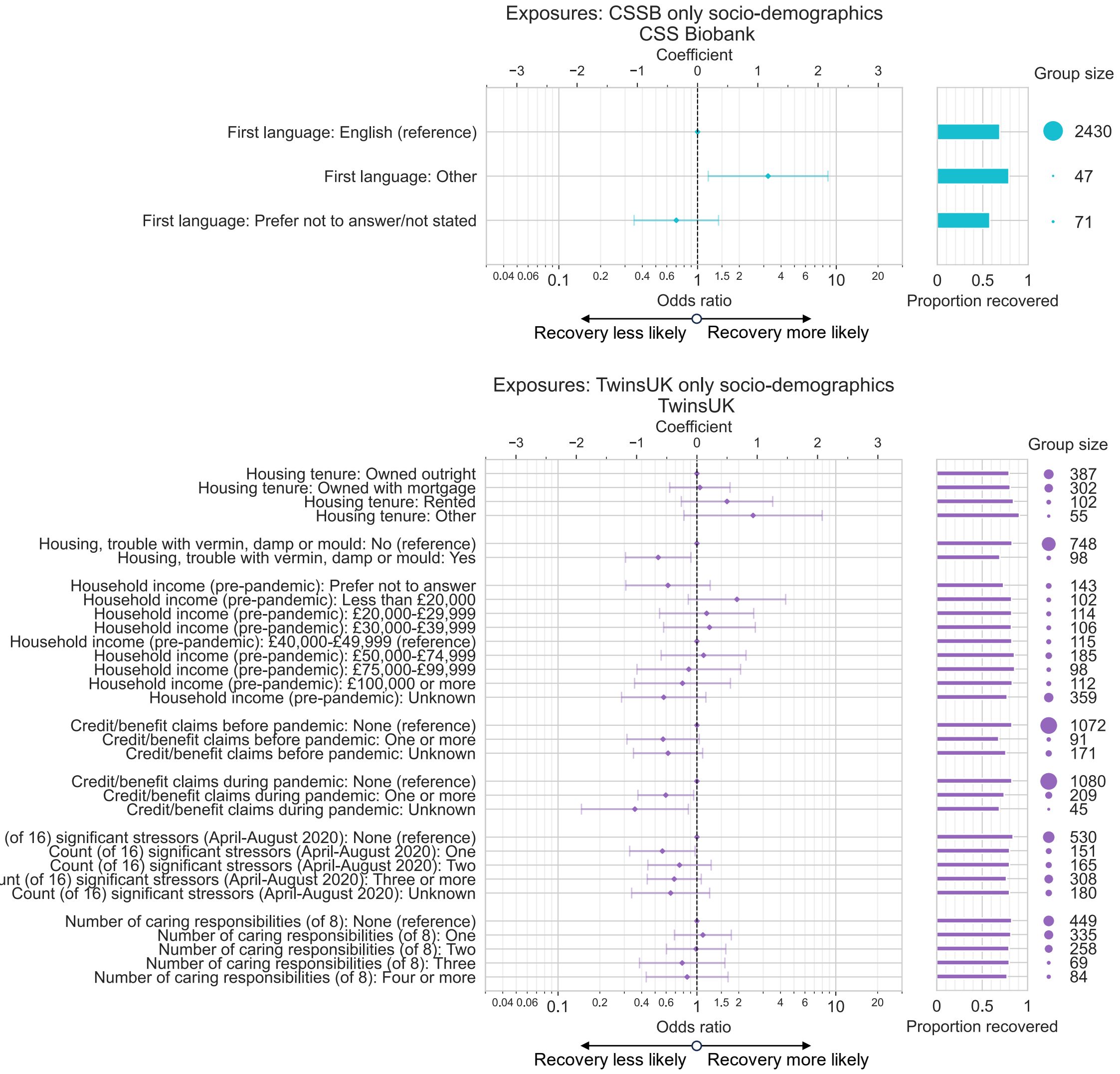

Figure S 8. **Associations between cohort-specific socio-demographics and recovery from COVID-19 in CSS Biobank and TwinsUK cohorts**. Odds ratio and 95% confidence intervals from logistic regression models testing association between recovery from COVID-19 and socio-demographic exposure variables unique to each cohort, among individuals with self-reported COVID-19 infection. Results for each exposure variable originate from distinct models, including age, sex, ethnic group, education, pre-pandemic socio-demographic factors and pre-pandemic health characteristics as potential confounding factors.
